# International recommendations for sleep and circadian research in aging and Alzheimer’s disease: a Delphi consensus study

**DOI:** 10.1101/2025.04.06.25325352

**Authors:** Claire André, Laura Stankeviciute, Johannes C Michaelian, Inga M Antonsdottir, Ruth M Benca, Elizabeth J Coulthard, Angela L D’Rozario, Derk-Jan Dijk, Sandra Gimenez, Maurizio Gorgoni, Yue Leng, Brendan P Lucey, Matthew P Pase, Stephanie R Rainey-Smith, Ivana Rosenzweig, Adam P Spira, Joseph R Winer, Géraldine Rauchs, Sharon L Naismith, Sleep and Circadian Rhythms Professional Interest Area of the International Society to Advance Alzheimer’s Research and Treatment (ISTAART)

## Abstract

**Introduction:** Research in the field of sleep, aging and dementia is rapidly growing. Consensus guidance is needed to facilitate high-quality research, comparability and consistency.

**Methods:** A modified Delphi consensus study was conducted by the *Sleep and Circadian Rhythms Professional Interest Area* of the International Society to Advance Alzheimer’s Research and Treatment (ISTAART) and an international panel of experts to establish recommendations for future research.

**Results:** After two voting rounds, the group determined by consensus (1) the most relevant sleep and circadian features to assess in the context of aging and dementia research, (2) established recommendations on data acquisition and report for future studies, and (3) identified high-priority future directions.

**Discussion:** This expert consensus study provides guidance to develop high-quality sleep and circadian research in the context of aging and dementia. Similar recommendations will need to be established for clinical practice.

## 1. Background

In recent years, research on sleep and circadian rhythms in the context of aging and dementia has grown exponentially. Sleep and circadian disturbances are highly prevalent among individuals with Alzheimer’s disease (AD) and related dementias, often manifesting years before diagnosis^1–5^. These disturbances include, but are not restricted to, shortened and more fragmented sleep, reduced slow wave sleep and rapid eye movement (REM) sleep, irregular sleep-wake patterns, and delayed sleep phase^1,4,6^.

Sleep-wake disturbances are often detectable before the onset of dementia, in individuals at greater risk of cognitive decline and are more pronounced in Mild Cognitive Impairment (MCI) compared to cognitively healthy older adults^2^. Emerging research suggests a bidirectional association between sleep and neurodegeneration, such that sleep disturbances may not only be a consequence of the degeneration of brain regions involved in sleep regulation^7^, but may also increase the vulnerability for, or exacerbate, Alzheimer’s and cerebrovascular disease pathologies that underpin cognitive decline and dementia^6–8^. Thus, it is plausible that early or timely interventions targeting sleep-wake disturbances may not only improve sleep and overall day-to-day cognition, mood and functioning, but may even mitigate disease processes, in turn delaying, slowing, or preventing cognitive decline.

The evidence supporting an etiological role of sleep-wake disturbance in the development of dementia is still evolving. Indeed, in the recent Lancet Commission report of modifiable dementia risk factors^9^, sleep was not formally classified as a modifiable risk factor. Although there has been a proliferation of research over the last decade, the field is hindered by important methodological factors, increasing the difficulty of synthesizing and drawing conclusions. Specifically, a variety of sleep disorders (e.g., obstructive sleep apnea, insomnia) and disturbances (e.g., poor self-reported sleep quality, both short and long sleep duration, sleep fragmentation) can be measured. Yet, their specific nuances and phenotype are often not fully described nor taken into account. In addition, a diverse range of methods and techniques are utilized in research settings to characterize sleep disturbance, ranging from self- or informant questionnaires and sleep diaries to more objective measures of rest-activity rhythms and sleep, measured via actigraphy, electroencephalography (EEG) and full polysomnography (PSG) respectively. Within these methods, considerable heterogeneity also exists in terms of measurement and analysis. Furthermore, different methods may be utilized according to the extent of cognitive impairment (i.e., cognitively intact *vs.* MCI or dementia), with a relative paucity of longitudinal studies. Recent years have also witnessed a rapid growth of newly-developed ‘wearable’ and ‘nearable’ devices, each of which would ideally undergo validation in the target population. Overall, these factors complicate the comparability between studies, synthesis within the literature and the application of findings into clinical settings.

In this context, the *Sleep and Circadian Rhythms Professional Interest Area* of the *International Society to Advance Alzheimer’s Research and Treatment* (ISTAART) conducted an expert consensus study to harmonize future research efforts. First, we aimed to establish research recommendations on (i) the most important data to acquire in future sleep and circadian rhythms studies in the context of aging and Alzheimer’s disease, (ii) the optimal methodological approaches to adopt in older populations with and without cognitive deficits, and (iii) the use of newly developed wearable and nearable devices. Second, we aimed to determine high-priority future directions for research, including which knowledge gaps should be addressed, and which sleep and circadian features, populations and designs should be prioritized.

## 2. Methods

### 2.1. Steering committee

A steering committee (SC) composed of experts in the field of sleep in aging and dementia was constituted between December 2022 and September 2023 by invitation of the chair (Sharon Naismith; chair of the ISTAART Sleep and Circadian Rhythms Professional Interest Area 2022-2024) and co-chair (Claire André; executive committee member of the ISTAART Sleep and Circadian Rhythms Professional Interest Area 2020-2024). The chair and co-chair invited members of the executive committee of the ISTAART Sleep and Circadian Rhythms Professional Interest Area (PIA) (n=3; L.S., G.R., I.M.A.) as well as specific international academic experts in the field of sleep, aging and dementia from different geographic regions (North America, Europe, Asia-Pacific) to be part of the SC. The final 18 voting SC members included: Sharon L. Naismith (chair; Australia), Claire André (co-chair; Canada), Laura Stankeviciute (Spain), Géraldine Rauchs (France), Inga M. Antonsdottir (U.S.A), Adam P. Spira (U.S.A), Brendan P. Lucey (U.S.A), Ruth M. Benca (U.S.A), Yue Leng (U.S.A), Joseph R. Winer (U.S.A), Angela L. D’Rozario (Australia), Stephanie R. Rainey-Smith (Australia), Matthew P. Pase (Australia), Derk-Jan Dijk (U.K.), Ivana Rosenzweig (U.K.), Elizabeth J. Coulthard (U.K.), Maurizio Gorgoni (Italy) and Sandra Gimenez (Spain). The SC also included Johannes C. Michaelian (Australia) as an early career researcher and key facilitator of ethical approvals, survey design and data quality. The terms of reference for the SC were to provide input into the aims, design and methods of the study, to participate in data collection in their capacity of international field experts, and to assist in the synthesis and interpretation of results. No funding was provided to SC members nor to the participants of the study.

### 2.2. Delphi process

A modified Delphi consensus approach was used to reach consensus and establish recommendations for future research. The study was granted ethics approval by the University of Sydney Human Research Ethics Committee (HREC) in October 2023 (project n°2023/538). The overall design of the Delphi process is summarized in **Figure 1**. The SC designed the questions and outcomes of the initial survey **(Appendix A)**, organized around three main axes:

1. Determine the most relevant sleep and circadian features in the context of aging and dementia;
2. Establish recommendations on data acquisition and reporting; and,
3. Identify high-priority future directions in the field of sleep, aging and dementia.

**Figure 1.**
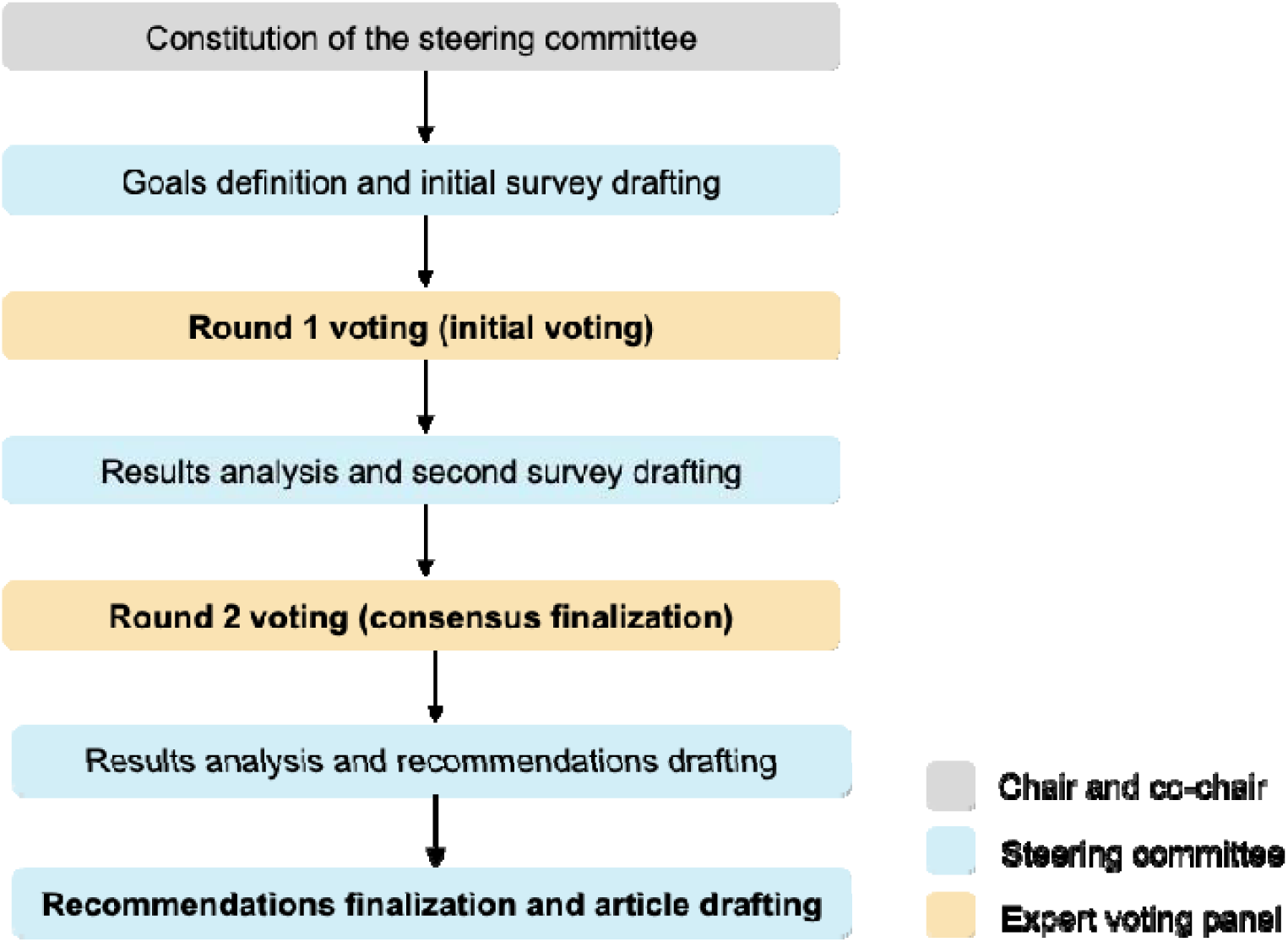
Summary of the Delphi consensus process.

The survey requested input regarding research recommendations (sought from all respondents) as well as recommendations for clinical settings (medical physicians or allied health). Here, we only report results related to research recommendations. Clinical recommendations will be reported separately.

Following SC input on a draft survey, further input on the content and wording of the draft was sought via a 1-hour workshop held at the 2023 Alzheimer’s Association International Conference in Amsterdam, Netherlands, gathering 23 SC and ISTAART members. After incorporating all feedback, the finalized survey (Delphi Round 1) was transferred to the Qualtrics XM platform (www.qualtrics.com) by J.C.M., I.M.A. and C.A., and disseminated online through the ISTAART Sleep and Circadian Rhythms PIA mailing list, to reach a broad audience of experts. The vote was open between November 2023 and February 2024 to all members (n∼480) of the ISTAART Sleep and Circadian Rhythms PIA and non-member researchers in the field of sleep, aging and dementia. Anonymous outputs were extracted and analyzed using SPSS (version 29). For each question, a consensus was considered achieved when 70% or more of participants voted for the same answer. During a virtual meeting, results were presented by the chair and co-chair to SC members, who provided feedback and suggestions for restructuring the design and content of a refined second-round survey **(Appendix B)**. Questions that reached consensus in Round 1 were not repeated in Round 2, unless the SC deemed the responses ambiguous or needing clarity. Questions that did not reach consensus in Round 1 were either sent back identically in Round 2, or refined following the SC recommendations. Round 2 online survey was implemented in Qualtrics and sent to participants who agreed to participate in May 2024. In July 2024, results were presented to the SC during a virtual meeting, and final recommendations were discussed with SC members during the 2024 Alzheimer’s Association International Conference in Philadelphia, USA, and were then approved by all SC members.

## 3. Results

### 3.1. Voting panel overview

During the first round of the Delphi process, 40 complete responses from 12 countries were received (United States of America: n=11, Australia: n=7, United Kingdom: n=5, Spain: n=5, Canada: n=2, France: n=2, Italy: n=2, Netherlands: n=2, Switzerland: n=1, Ireland: n=1, China: n=1, Iran: n=1). Of the sample, 60% identified as women and 40% as men, 7.5% indicated being from a low- or middle-income country, 82.5% identified as White, 12.5% identified as Asian, and 2.5% identified as Black.

In terms of the primary working environment, 52.5% indicated working in a university, 25% in a research center or institute, 12.5% in a hospital or health service, 7.5% in medical school, and 2.5% in industry. All 40 participants had research experience (including 95% actively conducting research), and 42.5% were early-career researchers (defined as <8 years post-PhD). Years of experience in research ranged between 1 and 45, with 15 participants (37.5%) having less than 10 years of experience, 16 (40%) having 10 to 19 years of experience, and 9 (22.5%) participants holding 20 or more years of experience.

With regards to research expertise, 24 indicated having experience in sleep medicine, 21 in sleep neurophysiology, 26 in Alzheimer’s and dementia biomarkers (n=21 with expertise in neuroimaging, n=19 with expertise on fluid biomarkers, n=6 with expertise on genetic biomarkers), 8 in epidemiology and public health, 5 in health services, 4 in basic science, 12 in clinical or applied research, 16 in clinical trials, 1 in engineering and/or computer science, and 2 in data science and/or artificial intelligence. Three participants worked in industry (sleep technology company or consulting).

Twenty-one participants had a clinical background (n=10 Neurologists, n=7 Sleep Physicians, n=3 Nurse Practitioners, n=3 Psychiatrists, n=2 Neurophysiologists, n=1 General Practitioner, n=1 Exercise Physiologist, n=1 Neuropsychologist, n=1 Clinical Psychologist, n=1 General Psychologist, n=1 not specified). Years of clinical experience ranged between 1 and 40 (n=9 between 1 and 10 years, n=7 between 11 and 20 years, n=4 between 21 and 30 years, and n=1 with 40 years of experience), and 17 were still active in clinical practice. Eleven participants (i.e., 27.5% of the panel) reported conflicts of interest, which are fully disclosed in the **Methods Supplement**.

Thirty-eight participants agreed to be contacted again for the second Delphi round, and we received a total of 32 complete responses to the second Delphi survey. Demographic characteristics of the voting panel for the second round were largely the same as the first round, and are fully described in the **Methods Supplement**.

### 3.2. Sleep and circadian features relevant to pathological ageing

In the first Delphi round, participants were asked to identify which sleep and circadian features were the most sensitive to preclinical AD, MCI or AD dementia, compared to normal aging. A large set of traditional sleep and circadian rhythms features and/or family of measures were presented (n=37, including 13 derived from clinical and self-reported data, 17 derived from PSG, and 7 derived from actigraphy recordings). Participants were asked to rate on a 4-point scale (strongly disagreed, disagreed, agreed or strongly agreed) whether the sleep and circadian features are sensitive to preclinical AD, MCI or AD dementia. The panel agreed on the relevance of 17 items for preclinical AD **(Supplementary Table 1)**, 29 items for MCI **(Supplementary Table 2)** and 27 items for AD dementia **(Supplementary Table 3)** (i.e., consensus was reached with at least 70% of the panel voting for the « agreed » or « strongly agreed » options).

In the second Delphi round, participants were asked to rate from the specific features identified as relevant by consensus in the first round, the 10 most suitable measures for each clinical category. These 10 best features are fully displayed in **Supplementary Table 4**. Based on the results, 11 measures were judged as the most relevant to assess in research examining pathological aging, most of them being recommended in all three clinical groups (i.e., preclinical AD, MCI and AD dementia), with no clear specificity between each clinical group **(Box 1)**.

#### Box 1 Measures rated as most relevant to assess in preclinical AD, MCI and AD dementia.

*Measures derived from objective sleep and circadian rhythm recordings (either actigraphy and/or PSG, when appropriate):*

- Sleep fragmentation indices (e.g., number of awakenings, arousals, stage shifts)
- Sleep efficiency
- Wake after sleep onset
- Total sleep time
- Duration or percentage spent in each sleep stage
- Non-rapid eye movement (NREM) sleep spectral power (including slow wave activity) and slow wave characteristics derived from PSG
- Rapid eye movement (REM) sleep spectral power (including REM EEG slowing)
- Daytime activity, including potential napping

*Measures derived from clinical interviews or self-reports:*

- General sleep disturbance and poor sleep quality
- Insomnia
- Excessive daytime sleepiness symptoms

### 3.3. Recommendations on data acquisition and reporting

#### 3.3.1. Best methods to assess sleep and circadian features

The panel was asked about which methods (i.e., informant self-report, patient self-report, actigraphy/accelerometer devices, in-lab PSG/EEG, at-home PSG/EEG) should be used to assess specific sleep and circadian features in the context of aging and dementia research. The results of round 1 showed a preference for PSG/EEG **(Supplementary Table 5)** but without a clear consensus towards in-lab or at-home settings. In round 2, both in-lab and at-home PSG/EEG options were combined, and the preference for in-lab or at-home settings was assessed in a separate question for clarity. In preclinical AD and MCI, PSG/EEG was selected as the best method to assess total sleep time, sleep onset latency, sleep efficiency, sleep fragmentation and wake after sleep onset, with actigraphy being the second-best choice **(Supplementary Table 5)**. In AD dementia, PSG/EEG was still the first option with actigraphy being the second-best choice for sleep onset latency, sleep efficiency, sleep fragmentation and wake after sleep onset, but with lower percentages in favor of PSG/EEG compared to actigraphy **(Supplementary Table 5)**. Both actigraphy and PSG/EEG were selected as equally optimal for assessing sleep duration/total sleep time in AD dementia. To assess daytime napping, actigraphy was chosen as the best method across all clinical categories (preclinical AD: 66.7%, MCI: 72.4%, AD dementia: 66.7%). Patient self-report was the second-best option for preclinical AD, and informant self-report was the second-best option for MCI and AD dementia **(Supplementary Table 5)**.

#### 3.3.2. Consensus on the minimum standards for research data collection and analysis in future studies and clinical trials

Regarding data collection, 77.5% of the panel declared that they routinely collected sleep data, including actigraphy (62.5%), self-report or clinical data (60%) and PSG/EEG (60%).

##### 3.3.2.1. Recruitment and characterization of participants

The panel was asked to select which participants characteristics need to be assessed and reported in addition to age and biological sex in sleep and/or circadian research studies in the context of aging and dementia. They were presented with 66 features including 24 demographics, clinical and biological features, 13 comorbidities and/or medical features, 10 sleep-related features, and 19 categories of medication. They were asked to select whether each feature was absolutely essential to assess, necessary to assess for a well-defined study, or not essential/optional to assess.

In the first Delphi round, a set of 52 features were deemed necessary to assess (i.e., when at least 70% of the votes included the combined options « absolutely essential » or « necessary for a well-defined study), including 15 demographic, clinical and biological items, 12 comorbidities, 8 sleep-related features, and 17 medication classes (see **Supplementary Table 6** for the full results).

In the second round, the panel was asked to select the five most essential measures to assess for each category, which are fully displayed in **Supplementary Table 7**. Regarding medication, participants were also asked to choose the optimal time frame within which sleep and/or psychotropic medication use should be assessed when designing clinical trials. The majority selected 1 month (n=9/21; 42.9%), followed by 2 weeks (n=7/21; 33.3%) and 1 week (n=2/21; 9.5%).

##### 3.3.2.2. Questionnaires and self-report data

We asked the panel about their confidence regarding self-reported sleep data in the context of aging and dementia **(Supplementary Table 8)**. The results suggested very low to no confidence in the reliability of data derived from single-item questions about sleep (i.e., 76.3% voted for the “not confident at all” or “not confident” options), with 100% declaring that the quality of this type of data depends on the cognitive status of the participant **(Supplementary Table 8)**. Regarding informant-related data, 60.5% of the panel reported being not confident in their reliability, with 97.4% judging that the quality depends on the cognitive status of the informant **(Supplementary Table 8)**. Another limitation is the absence of sleep questionnaires specifically designed for older populations: 92.1% of the panel reported that sleep questionnaires should be re-validated for older populations, patients with MCI or AD dementia, and 94.7% reported that the scoring of existing questionnaires should be adapted or standardized based on these specific populations **(Supplementary Table 8)**.

##### 3.3.2.3. Actigraphy and accelerometer-based wearable devices

For descriptive purposes, the first Delphi round asked the panel about their experience with actigraphy and accelerometer wearable devices. Seventy-five percent of the panel (n=30) reported using actigraphy and other research-quality accelerometer wearable devices (e.g., Actigraph, GENEActiv, etc.), 15% (n=6) reported using commercially-available accelerometer wearable devices (e.g., Fitbit, Apple Watch, etc.), and 10% (n=4) reported using none of these devices in their research.

###### Actigraphy recording duration

The majority of the panel stated that the *ideal duration* of actigraphy recordings to assess sleep and circadian rest-activity rhythms was ≥14 days (67.7% and 77.4% respectively) **(Supplementary Table 9)**. Additionally, the *minimum duration* for actigraphy-based sleep and circadian recording was deemed to be 7 to 13 days (71% and 61.3% respectively) **(Supplementary Table 9)**.

###### Actigraphy data collection

The large majority (86.8%) of the panel voted in favor of always using a sleep diary in conjunction with actigraphy, which would notably probe for the following key factors: sleep latency, sleep timing, daytime napping, the use of caffeine, alcohol or medication use, feeling refreshed in the morning, sleep disruption and associated symptoms **(Supplementary Table 9)**.

###### Actigraphy data processing

The panel was mostly (66.7%) confident about using sleep diary data as a tool for interpreting actigraphy recordings, and using it for quality control purposes, while 33.3% were not confident. Most of the panel (61.8%) sometimes change actigraphy recordings/outputs based on sleep diary reports, while almost one third (32.4%) do this often **(Supplementary Table 9)**. The majority (92.9%) of the panel recommended the use of both in-house and software-implemented algorithms to process actigraphy recordings **(Supplementary Table 9)**.

##### 3.3.2.4. Polysomnography

The panel was first asked about which electrode locations need to be included as a priority in research settings. Electrode sites receiving more than 40% of votes were F3, F4, C3, C4, O1 and O2, followed by Cz (37.5%), Fz (30%), Pz (27.5%), with P3, P4, Fp1 and Fp2 all receiving 22.5% of the votes **(Supplementary Table 10)**. Other electrodes (i.e., T3, T4, T5, T6, F7, F8) were endorsed by less than 20% of the panel.

When asked about their preference for in-lab or at-home PSG/EEG settings, there was no clear consensus, but almost half of the panel judged that both were equally acceptable in preclinical AD (48.6%) and MCI (45.5%), while a preference for at-home PSG/EEG assessment was expressed for AD dementia patients (57.6%) **(Supplementary Table 11)**. However, in the second round, the majority (86.5%) of the panel agreed that for optimal compromise between feasibility and ecological validity, opting for in-lab or at-home EEG/PSG still depends on both the research question and the clinical group.

#### 3.3.3. Confidence in the use of newly available wearable and nearable devices

In recent years, the development and use of wearable and nearable devices for sleep diagnostics and monitoring have increased. These potentially offer distinct advantages over PSG in terms of ecological validity, longer recording periods, lower costs, and easier set-up and administration. It is noted that such devices are not yet used routinely in clinical practice, and that end-user engagement is critical in testing and utilizing such devices across the spectrum of pathological aging, particularly if translation is to be a core aim of the research.

Research-quality devices for sleep assessment are engineered to deliver highly precise and reliable data (e.g., accelerometry, heart-rate variability, temperature, sleep staging, EEG features) essential for scientific and clinical investigations. These devices undergo rigorous validation and conform to stringent regulatory standards, such as obtaining a medical device CE mark and FDA approvals. They ideally provide access to raw data, enabling researchers to perform detailed analysis and customization in research applications.

Consumer-quality devices, on the other hand, prioritize ease of use, convenience, and affordability. Such devices typically offer patented/undisclosed algorithm-derived outputs with limited access to raw data, prioritizing user-friendly accessible insights into sleep patterns (i.e., sleep/restfulness scores) over the depth of analysis possible with research-grade equipment. In addition, consumer-quality devices do not necessarily follow the medical-approval pathway, often bypassing the stringent validation and regulatory processes required for medical-grade qualifications and clinical applications. Despite these limitations, there is potential research value in consumer-quality devices, as they may already be successfully used by the participants for personal health monitoring. It is important to acknowledge that the distinction between research and consumer-quality devices is not always clear-cut, with significant overlap between the two categories. However, for the purpose of this paper, “research-quality” will refer to medical-grade and validated devices, while “consumer-quality” will define devices that are commercially available but not approved as medical devices by regulatory authorities.

##### 3.3.3.1. Accelerometer devices

In the first Delphi round, the panel expressed confidence in the use and validity of accelerometer devices recording movement only (73.5% confident, 26.5% not confident), and devices recording movement combined with other types of data (e.g., heart rate, O2) (78.8% confident, 21.2% not confident) **(Supplementary Table 12)**. When distinguishing research-quality and consumer-quality devices, the panel was confident in the validity of research-quality devices in cognitively unimpaired older adults (94.4% confident, 5.6% not confident), as well as in patients with MCI or AD dementia (80% confident, 20% not confident) **(Supplementary Table 12)**. However, the majority of panel expressed a lack of confidence in the validity of consumer-quality devices across all groups (cognitively unimpaired older adults [71.4% non-confident, 28.6% confident], MCI or AD dementia [81.8% non-confident, 18.2% confident]) **(Supplementary Table 12)**.

During the second Delphi round, we asked more precisely about the panel’s confidence in research-quality versus consumer-quality accelerometer devices recording movement only for measuring specific sleep and circadian features. There was a consensus for confidence in research-quality accelerometer devices for measuring total sleep time, sleep onset and offset, sleep efficiency, wake after sleep onset, daytime activity (e.g., napping) and circadian features **(Supplementary Table 13)**, but a trend for non-confidence regarding the assessment of sleep onset latency. However, the panel globally expressed a lack of confidence in consumer-quality devices for measuring all of these parameters (i.e., above the 70% consensus threshold for sleep onset latency, sleep onset and offset, sleep efficiency and daytime activity, and a similar trend for total sleep time, wake after sleep onset and circadian features) **(Supplementary Table 13)**.

##### 3.3.3.2. EEG-based devices

The panel was confident in the validity of such devices in cognitively unimpaired individuals (79.3% confident, 20.7% not confident), while there was no clear confidence in patients with MCI or AD (65.5% confident, 34.4% not confident) **(Supplementary Table 12)**. When distinguishing research-quality and consumer-quality devices in MCI and AD patients during the second Delphi round, the panel was confident in the validity of research-quality devices (confident: 71%, not confident: 29%) but not consumer-quality devices (confident: 13.3%, not confident: 86.7%) **(Supplementary Table 14)**. The panel was overall mixed regarding the validity of new EEG-based wearables to replace full PSG recordings (44.9% confident, 55.1% not confident) on the first Delphi round **(Supplementary Table 12)**, with 85.3% highlighting that such devices should be combined with other devices assessing complementary aspects (e.g., heart rate, O2, etc.) **(Supplementary Table 12)**. In the second Delphi round, we distinguished research-quality and consumer-quality devices, and the votes were still mixed for research-quality devices (confident: 43.3%, not confident: 56.7%). There was a clear non-confidence for consumer-quality EEG devices (confident: 16.7%, not confident: 83.3%) **(Supplementary Table 14)**. However, the panel was confident in using oximeters at home to assess OSA in cognitively unimpaired older adults (confident: 83.3%, not confident: 16.7%) **(Supplementary Table 12)**.

### 3.4. Moving the field forward: high-priority future directions

#### 3.4.1. What further evidence is needed to establish or discard sleep problems as a risk factor for dementia?

We asked the panel to identify the five most critical lines of evidence required to establish or discard sleep problems as a risk factor for dementia. In Delphi round 1, the five options receiving the most votes were: more studies with longitudinal follow-up (90%), more robust clinical trial data on the effect of sleep treatments on cognition and/or AD biomarkers (80%), more continuous monitoring of sleep (e.g., several days of recording in a row) (70%), more robust clinical trial data on the effect of sleep treatments on dementia onset/prevention (70%), more biomarker/basic science studies (62.5%) **(Table 1)**.

**Table 1.** Further evidence needed to establish or discard sleep problems as a risk factor for dementia.

| Answer options | n/40 | % | Rank |
| --- | --- | --- | --- |
| More studies with longitudinal follow-up | 36 | 90.0 | **1** |
| More robust clinical trial data on the effect of sleep treatments on cognition, AD biomarkers | 32 | 80.0 | **2** |
| More continuous monitoring of sleep (e.g., several days of recording in a row) | 28 | 70.0 | **3** |
| More robust clinical trial data on the effect of sleep treatments on dementia onset/prevention | 28 | 70.0 |  |
| More biomarker/basic science studies | 25 | 62.5 | **4** |
| More mechanistic EEG studies, for example examining sleep microarchitecture | 23 | 57.5 | **5** |
| More studies assessing sleep using PSG | 16 | 40.0 | 6 |
| More studies in clinical populations | 12 | 30.0 | 7 |
| More epidemiological studies | 9 | 22.5 | 8 |
| More studies using self-report questionnaires | 3 | 7.5 | 9 |

Results of the vote are expressed in terms of frequency (n/40), percentage (%) and rank. The five best-ranking options using a sum of frequencies approach are highlighted in bold.
*Abbreviations: AD, Alzheimer's disease; EEG, electroencephalography; PSG, polysomnography.*

#### 3.4.2. High priority sleep and circadian features warranting examination

The panel was also asked which aspects of sleep and circadian research need to be studied as a priority in future studies. In the first round, 20 items were selected as important for future studies by the panel: slow waves, sleep spindles, the harmonization of actigraphy methodologies, REM sleep, circadian rhythms using gold-standard circadian outputs, N3 sleep and slow wave sleep, quantitative EEG/spectral analyses, EEG connectivity and coupling, sleep fragmentation, OSA, general actigraphy features, actigraphy-defined sleep variability, insomnia, sleep duration, sleep cycles, N2 sleep, napping, K complexes, epilepsy spike characteristics, parasomnias/RBD **(Supplementary Table 15)**. In the second round, the panel were asked to refine this selection, which allowed the 10 best-ranking items to be identified using a sum of frequencies approach **(Table 2)**. High-priority features and directions for future research included circadian rhythms, obstructive sleep apnea, REM sleep, EEG connectivity and coupling measures, spectral analyses and quantitative EEG, the harmonization of actigraphy methodology, insomnia, napping, sleep fragmentation, sleep spindles, epilepsy spike characteristics, N3 sleep/slow wave sleep, and day-to-day variability of sleep patterns **(Table 2)**.

**Table 2.** High-priority aspects of sleep and circadian research – Delphi round 2.

| Feature | Most important and requires further research | Most understudied to date, yet potentially relevant | Frequency (n/32) | Rank |
| --- | --- | --- | --- | --- |
| Circadian rhythms | 11 | 13 | 24 | **1** |
| Obstructive sleep apnea | 18 | 3 | 21 | **2** |
| REM sleep | 10 | 9 | 19 | **3** |
| EEG connectivity/coupling | 6 | 12 | 18 | **4** |
| Spectral analyses/quantitative EEG | 9 | 9 | 18 |  |
| Harmonization of actigraphy methodologies | 3 | 12 | 15 | **5** |
| Insomnia | 12 | 3 | 15 |  |
| Naps | 8 | 6 | 14 | **6** |
| Sleep fragmentation | 8 | 6 | 14 |  |
| Sleep spindles | 7 | 6 | 13 | **7** |
| Epilepsy spike characteristics | 2 | 10 | 12 | **8** |
| N3 sleep / slow wave sleep | 10 | 1 | 11 | **9** |
| Actigraphy-defined sleep variability | 4 | 6 | 10 | **10** |
| K complexes | 4 | 5 | 9 | 11 |
| Sleep cycles | 3 | 6 | 9 | 11 |
| Slow waves | 6 | 3 | 9 | 11 |
| Sleep-related movement disorders (e.g., PLMD) | 1 | 7 | 8 | 12 |
| Excessive daytime sleepiness (e.g., ESS) | 2 | 5 | 7 | 13 |
| Hypersomnolence disorders | 2 | 5 | 7 | 13 |
| Parasomnias/RBD | 4 | 3 | 7 | 13 |
| Actigraphy features generally | 5 | 1 | 6 | 14 |
| Global self-reported measures (e.g., PSQI) | 4 | 2 | 6 | 14 |
| Sleep duration | 2 | 2 | 4 | 15 |
| N1 sleep | 0 | 0 | 0 | 16 |
| N2 sleep | 0 | 0 | 0 | 16 |

Results of the vote are expressed in terms of frequency (n/32), percentage (%) and rank. The 10 best-ranking options using a sum of frequencies approach are highlighted in bold. Abbreviations: EEG, electroencephalography; ESS, Epworth Sleepiness Scale; PLMD, periodic limb movement disorder; PSG, polysomnography; PSQI, Pittsburgh Sleep Quality Index; RBD, REM sleep behavior disorder; REM, rapid eye movement.

#### 3.4.3. High priority understudied populations warranting further research

We then aimed to seek consensus on the most understudied populations in sleep and circadian research, to inform potential future research priorities. The panel was presented with 13 options, and were asked to judge whether they were ‘not important’, ‘minimally important’, ‘moderately important’ or ‘strongly important’. During the first Delphi round, all items were judged as important **(Supplementary Table 16)**. Therefore, we refined the question in the second round, by asking the panel to select the five groups with the highest priority. Using a sum of frequency approach, the panel determined that studies presenting stratifications according to biological sex, individuals with shift work history, race/ethnic disparities, non-AD dementias and middle-aged groups (40-65 years old) were high priority **(Table 3)**.

**Table 3.** Identification of understudied populations – Delphi round 2.

| **Group** | **Frequency (n/32)** |
| --- | --- |
| Stratification according to biological sex | **20** |
| Individuals with shift work history | **14** |
| Race/ethnic disparities | **14** |
| Non-AD dementias | **14** |
| Middle aged groups (40-65 years old) | **14** |
| Extreme chronotypes | 13 |
| Individuals with sleep disorders | 11 |
| People in low- and middle-income countries | 11 |
| Individuals with genetic risk factors for AD (e.g., APOE ε4 carriers) | 10 |
| Individuals with mental health disorders | 10 |
| Long sleepers (>9h) | 5 |
| Short sleepers (<6h) | 4 |
| Older groups (>65 years old) | 2 |

Results of the vote are expressed in terms of frequency (n/32). The five groups with the highest priority as determined using a sum of frequencies approach are indicated in bold.
*Abbreviations: AD, Alzheimer's disease; APOE, apolipoprotein E; h, hour.*

#### 3.4.4. High priority future directions for the field

Finally, the panel was asked to determine high priority future research directions for the field of sleep and circadian rhythms in the context of aging and dementia. On the first Delphi round, we obtained a consensus, defined as at least 70% of the votes for the ‘strongly important’ option, on the importance of:

1. Gathering more evidence on the effect of treating sleep disturbances as a way to slow cognitive decline and/or impact the accumulation of AD pathology (strongly important: 86.49%);
2. Determining whether sleep interventions work in individuals at risk for AD (e.g., with risk factors or cognitive impairment) (strongly important: 86.49%);
3. Developing circadian treatment and interventions (strongly important: 80.56%);
4. Developing a global consortium for sleep and dementia data sharing (strongly important: 80.56%);
5. Gaining a better understanding of the functions of sleep (e.g., glymphatic clearance, cognition, etc.) (strongly important: 80.56%);
6. Determining if sleep disturbance should be screened for in dementia diagnosis (strongly important: 78.38%);
7. Determining if sleep disturbance should be screened for in people *at risk* for AD (e.g., preclinical and prodromal AD) (strongly important: 77.78%); and,
8. Identifying critical/optimal time windows within which sleep disturbances should be identified to optimize brain health (strongly important: 75%) **(Table 4)**.

**Table 4.** Identification of high-priority future directions in the field.

| Answer options |  | Not important | Minimally important | Moderately important | Strongly important | Total |
| --- | --- | --- | --- | --- | --- | --- |
| Study sleep in under-studied and under-represented populations (e.g., ethnic disparities, sex and gender differences, other types of dementias, etc.) | n | 0 | 2 | 11 | 22 | 35 |
|  | % | 0 | 5.71 | 31.43 | 62.86 | 100 |
| Re-validate existing sleep questionnaires and potentially adapt their scoring to older populations with cognitive impairment | n | 0 | 4 | 15 | 17 | 36 |
|  | % | 0 | 11.11 | 41.67 | 47.22 | 100 |
| Develop a new sleep screening questionnaire specifically adapted to older people with cognitive impairment | n | 0 | 2 | 13 | 21 | 36 |
|  | % | 0 | 5.56 | 36.11 | 58.33 | 100 |
| Establish normative sleep data for older populations (for architecture, and microstructure/qEEG). | n | 0 | 2 | 9 | 24 | 35 |
|  | % | 0 | 5.71 | 25.71 | 68.57 | 100 |
| Identify critical/optimal time windows within which sleep disturbance should be identified to optimize brain health | n | 0 | 0 | 9 | 27 | 36 |
|  | % | 0 | 0 | 25 | **75** | 100 |
| Better understand the functions of sleep (e.g., glymphatics, cognition...) | n | 0 | 0 | 7 | 29 | 36 |
|  | % | 0 | 0 | 19.44 | **80.56** | 100 |
| Study the impact of lifestyle factors (e.g., physical activity) on sleep. | n | 0 | 0 | 15 | 21 | 36 |
|  | % | 0 | 0 | 41.67 | 58.33 | 100 |
| Gather more evidence on the treatment of sleep disorders/disturbances as a way to slow cognitive decline and/or impact the accumulation of AD pathology. | n | 0 | 0 | 5 | 32 | 37 |
|  | % | 0 | 0 | 13.51 | **86.49** | 100 |
| Determine if sleep should be screened for in dementia diagnosis | n | 0 | 2 | 6 | 29 | 37 |
|  | % | 0 | 5.41 | 16.22 | **78.38** | 100 |
| Determine if sleep should be screened in people at risk for AD (e.g., preclinical and prodromal AD) | n | 0 | 0 | 8 | 28 | 36 |
|  | % | 0 | 0 | 22.22 | **77.78** | 100 |
| Determine whether sleep interventions work in people at risk for AD (e.g., with cognitive impairment, risk factors) | n | 0 | 0 | 5 | 32 | 37 |
|  | % | 0 | 0 | 13.51 | **86.49** | 100 |
| Determine the role of circadian misalignment in AD pathology | n | 0 | 1 | 10 | 25 | 36 |
|  | % | 0 | 2.78 | 27.78 | 69.44 | 100 |
| Develop circadian treatments | n | 0 | 0 | 7 | 29 | 36 |
|  | % | 0 | 0 | 19.44 | **80.56** | 100 |
| Develop a global consortium for sleep and dementia data sharing | n | 0 | 1 | 6 | 29 | 36 |
|  | % | 0 | 2.78 | 16.67 | **80.56** | 100 |

High priority future directions selected during the first Delphi round. Results are expressed in terms on frequency and percentage. Percentages highlighted in bold reached the 70% consensus threshold. *Abbreviations: AD, Alzheimer's disease; qEEG, quantitative electroencephalography.*

## 4. Discussion

This study integrates views and perspectives of global leaders in sleep and circadian rhythms as they pertain to aging, MCI and AD dementia, as well as stakeholders working in this field. As a core aim, it seeks to provide guidance to harmonize and synergize research to increase comparability and clarity within the field.

Final recommendations on data acquisition and reporting for future research are described in **Box 2**. They document the core (mandatory) and core-plus (strongly encouraged) data to document in future studies and clinical trials on sleep and circadian rhythms in the context of aging and dementia, and provide methodological recommendations while considering the availability of various methodologies and emerging wearable and nearable devices.

The panel also identified high-priority future research directions detailed in **Box 3**, which include prioritizing studies (i) on several highly-relevant aspects of sleep (e.g., obstructive sleep apnea, insomnia, slow wave sleep, REM sleep, sleep fragmentation, quantitative EEG and connectivity, night-to-night sleep variability, daytime napping) and circadian rhythms, especially in understudied populations (e.g., sex-stratified designs, race/ethnic disparities, individuals with shift work history, non-AD dementias and middle-aged individuals), (ii) increasing our understanding of sleep functions and the associations between sleep and neurodegenerative processes, and (iii) conducting clinical trials assessing the effect of sleep and circadian interventions for dementia prevention. In terms of methodological advances, the panel recommends the validation of existing tools specifically in older populations, the development of new tools specifically adapted to older populations, prioritizing longitudinal designs and more continuous assessments of sleep (e.g., multi-night measurements). Finally, the panel recommends establishing a global consortium focused on harmonizing existing data and facilitating the sharing of new findings on sleep and circadian rhythms in the context of aging and dementia.

This study is the first to address this complex field in a collaborative framework. The SC included representation from many global leaders, with participants from 12 diverse regions, including 7.5% from low- and middle-income countries. However, there were notable exceptions from South America, Asia, and Africa. It is noted that the target audience within the ISTAART Sleep and Circadian Rhythms PIA includes members from these regions, who were invited via email to participate, but more focused strategies may have been necessary to foster participation. Additional barriers to a more global participation may have included the fact that the survey invitation and content were in English and that there was a limited time window within which the survey was open for participation. We acknowledge that enabling a longer time period could have resulted in greater participation rates. Of note, we utilized a Delphi method, which is favored for its participatory approach to consensus through iterative voting and feedback. However, the traditional method was modified in order to strike a balance of survey length design and pragmatism. Finally, the findings and recommendations presented here are only intended for research settings, and are not intended to be a guide for clinical practice, which will be reported in a subsequent publication.

#### Box 2 Recommendations on data acquisition and reporting for future research.

1. **Core data to document in future studies and clinical trials (mandatory):**

- <u>Demographic, clinical and biological features</u>: cognitive diagnosis, education level and body mass index.
- <u>Comorbidities and medical history</u>: neurological diseases, depressive symptoms, psychiatric disorders, cardiovascular diseases and alcohol intake.
- <u>Sleep and circadian-related features</u>: insomnia symptoms, OSA diagnosis, OSA treatment, current shift work and restless legs syndrome.
- <u>Medication</u>: benzodiazepines, non-benzodiazepine receptor agonist hypnotics, antidepressants, other approved hypnotics (e.g., DORAs, ramelteon), Trazodone and antipsychotics.
2. **Core-plus data to document (strongly recommended if available):**

- Biomarker profile (e.g., amyloid/tau/neurodegeneration).
- *APOE ε4* status.
3. **Methodological recommendations for the assessment of sleep and circadian rhythms in the context of aging and dementia:**

- Favor objective methods to assess sleep and circadian rhythms in older populations:

- PSG/EEG should be prioritized for standard sleep parameters. Alternatively, the use of actigraphy is recommended.
- Actigraphy is recommended to assess daytime napping. Alternatively, self-reported in preclinical AD, and informant report in MCI/AD dementia are recommended.
- Both in-lab and at-home PSG/EEG settings are acceptable in preclinical AD and MCI, but at-home PSG/EEG is preferred in patients with dementia.
- Avoid actigraphy recordings of less than 7 days. The recommended minimum standard duration is 7-13 days, while the recommended ideal duration is ≥14 days.
- It is recommended to always propose a sleep diary in conjunction with actigraphy to aid data interpretation and processing.
- Avoid single-item self-reported data to document sleep disturbances.
- Ensure the cognitive status of informants when analyzing informant-reported sleep data.
- Interpret data derived from consumer-quality wearable and nearable devices with caution.

### Box 3 High-priority future directions for the field.

**1) Type of research to prioritize:**

⍰ <u>Research on high-priority sleep and circadian aspects:</u> Measures of circadian rhythms, obstructive sleep apnea, REM sleep, EEG connectivity/coupling, spectral analyses and quantitative EEG, harmonization of actigraphy methodologies, insomnia, daytime napping, sleep fragmentation, sleep spindles, epilepsy spikes, N3 sleep and slow waves, night-to-night sleep variability.
⍰ Research aiming at better understanding the functions of sleep (e.g., glymphatic clearance, cognition), its associations with AD pathology and other potential pathological pathways (e.g., vascular, neuroinflammation).
⍰ <u>Research in understudied populations</u>: sex-stratified studies, individuals with shift work history, race/ethnic disparities, non-AD dementias and middle-aged groups (40-65 years old).
⍰ Clinical trials assessing the effect of sleep and circadian interventions as tools to prevent dementia.

- Determine if sleep interventions are feasible and effective in individuals at-risk for AD.
- Develop circadian interventions suited for older individuals with and without cognitive deficits.
- Gather more evidence on the effect of the treatment of sleep disturbances as a way to slow cognitive decline and/or impact the accumulation of AD pathology.
- Determine the optimal time windows within which sleep disturbances should be screened and treated to promote brain health.
⍰ Research aiming at determining the added value of sleep disturbances diagnosis in AD detection and diagnosis at different stages of the disease (dementia, prodromal and preclinical stages).

**2) Foster methodological advances:**

⍰ Validate existing sleep and circadian assessment tools specifically in older populations, and develop new tools adapted for older participants:

- Re-validate existing sleep questionnaires specifically in older populations and/or develop a geriatric sleep questionnaire.
- Validate newly-available research-quality and consumer-quality wearable and nearable devices specifically in older populations with and without cognitive deficits.
⍰ Prioritize longitudinal study designs.
⍰ Adopt more continuous and multi-night assessments of sleep.

**3) Create a global consortium to share new data on sleep in the context of aging and dementia, and harmonize existing data.**

## Acknowledgements, funding and disclosures

### a) Consent Statement

All human subjects provided informed consent.

## Data Availability

All data produced in the present study are available upon reasonable request to the authors

## Acknowledgements

This manuscript was facilitated by the Alzheimer’s Association International Society to Advance Alzheimer’s Research and Treatment (ISTAART), through the Sleep and Circadian Rhythms professional interest area (PIA). The views and opinions expressed by authors in this publication represent those of the authors and do not necessarily reflect those of the PIA membership, ISTAART or the Alzheimer’s Association. The authors would like to thank all the participants of the voting panel, as well as Jodi Titiner (ISTAART staff).

## 5. Keywords

Dementia, MCI, sleep, circadian rhythms, Delphi, consensus, actigraphy, polysomnography, wearables, questionnaires, recommendations

