## Supplementary for "International recommendations for sleep and circadian research in aging and Alzheimer’s disease: a Delphi consensus study"

**Methods Supplement: Conflict of interest of the voting panel.**

**Methods Supplement: Demographic characteristics of the voting panel – Delphi round 2.**

**Supplementary Table 1. Sleep and circadian features sensitive to preclinical AD – Delphi round 1.**

**Supplementary Table 2. Sleep and circadian features sensitive to MCI – Delphi round 1.**

**Supplementary Table 3. Sleep and circadian features sensitive to AD dementia – Delphi round 1.**

**Supplementary Table 4: Sleep and circadian features sensitive to pathological aging – Delphi round 2.**

**Supplementary Table 5. Best method to assess sleep and circadian features.**

**Supplementary Table 6. Minimum standard for data collection – Delphi round 1.**

**Supplementary Table 7. Minimum standard for data collection – Delphi round 2.**

**Supplementary Table 8. Confidence in self-reported sleep data.**

**Supplementary Table 9. Confidence and recommendations for actigraphy data.**

**Supplementary Table 10. Recommendations for polysomnography and EEG data.**

**Supplementary Table 11. Preference for in-lab or at-home PSG/EEG settings.**

**Supplementary Table 12. Confidence in the use of newly available wearable and nearable devices.**

**Supplementary Table 13. Confidence in new movement-based wearables and nearables for measuring specific sleep and circadian features in a research context.**

**Supplementary Table 14. Confidence in new EEG-based wearables – Delphi round 2.**

**Supplementary Table 15. Identification of understudied sleep and circadian features – Delphi round 1.**

**Supplementary Table 16. Identification of understudied populations – Delphi round 1.**

**Methods Supplement: Conflict of interest of the voting panel.**

Twenty-nine experts (72.5% of the panel) declared having no conflict of interest to disclose. The remaining 11 experts (27.5% of the panel) self-reported the following potential conflict of interest:

| **Description** | **Frequency (n)** |
| --- | --- |
| *Consultancies* | *7* |
| “Eisai, Idorsia, Haleon, Jazz, Merck” | 1 |
| “Eisai, Roche” | 1 |
| “Eli Lilly, Beacon Biosignals, OrbiMed, GLG Group” | 1 |
| “Merck, BellSant, Inc., Sequoia Neurovitality” | 1 |
| “Paladin Labs, Jazz Pharmaceuticals, Eisai, Idorsia” | 1 |
| “Pharma companies” | 1 |
| *Honoraria* | *2* |
| “Nutricia” | 1 |
| “Paladin Labs, Jazz Pharmaceuticals, Eisai, Idorsia” | 1 |
| *Employment in industry* | *1* |
| “Employee of the PHARMO Institute for Drug Outcomes Research” | 1 |
| *Other (please specify)* | *2* |
| “Received equipment for free from technology company” | 1 |
| “Research support” | 1 |

**Methods Supplement: Demographic characteristics of the voting panel – Delphi round 2.**

On the second round of the Delphi process, 32 complete responses from 10 countries were received (United States of America: n=8, United Kingdom: n=6, Australia: n=5, Spain: n=4, Canada: n=2, Italy: n=2, The Netherlands: n=2, France: n=1, Switzerland: n=1, China: n=1). The profile of the panel was similar to round 1. In terms of demographics, 56.3% of the participants identified as women and 40.6% as men while 1 preferred not to answer (3.1%); 3.1% indicated being from a low- or middle-income country; 84.4% identified as White, 12.5% identified as Asian, 3.1% identified as Black/African American. In terms of primary working environment, 33.3% indicated working in a research center or institute, 27.3% in a university, 18.2% in a hospital or health service, 15.2% in medical school, and 3% in the industry. All participants were still actively conducting research, and 13 (42%) were early-career researchers (defined as <8 years post-PhD). Years of experience in research ranged between 1 and 45, with 11 presenting with less than 10 years of experience, 12 with 10 to 19 years of experience, and 7 with 20 or more years of experience (2 did not specify years of experience). In terms of research expertise, 24 indicated having experience in Alzheimer’s and dementia biomarkers, 20 in clinical or applied research, 19 in sleep neurophysiology, 16 in sleep medicine, 15 in clinical trials, 7 in epidemiology and public health, 5 in basic science, 4 in data science and/or artificial intelligence, 3 in health services, 1 in computer science and/or engineering, and 1 worked in industry (sleep technology company). Twenty-two participants also reported having a clinical background (n=9 Neurologists, n=6 Sleep Physicians, n=3 Psychiatrists, n=2 Nurse Practitioners, n=1 Neuropsychologist, n=1 Exercise Physiologist). Years of clinical experience ranged between 3 and 40 (n=3 between 1 and 9 years, n=6 between 10 and 19 years, n=6 with at least 20 years of experience), and 13 were still in activity.

**Supplementary Table 1. Sleep and circadian features sensitive to preclinical AD – Delphi round 1.**

| **Variable** | | **Strongly Disagree** | **Disagree** | **Agree** | **Strongly Agree** | **Disagree (total)** | **Agree (total)** | **Total** |
| --- | --- | --- | --- | --- | --- | --- | --- | --- |
| **CLINICAL AND SELF-REPORTED DATA** | | | | | | | | |
| General sleep disturbance/poor sleep quality | n | 0 | 10 | 13 | 15 | 10 | 28 | 38 |
|  | % | 0 | 26.3 | 34.2 | 39.5 | 26.3 | **73.7** | 100 |
| Excessive daytime sleepiness | n | 4 | 10 | 14 | 5 | 14 | 19 | 33 |
|  | % | 12.1 | 30.3 | 42.4 | 15.2 | 42.4 | 57.6 | 100 |
| Sleep-related breathing disorders (e.g., OSA) symptoms | n | 3 | 9 | 13 | 10 | 12 | 23 | 35 |
|  | % | 8.6 | 25.7 | 37.1 | 28.6 | 34.3 | 65.7 | 100 |
| Parasomnias (e.g., RBD) and related symptoms | n | 7 | 13 | 6 | 3 | 20 | 9 | 29 |
|  | % | 24.1 | 44.8 | 20.7 | 10.3 | 68.9 | 31 | 100 |
| Sleep-related movement disorders (e.g., PLMD) | n | 6 | 15 | 6 | 1 | 21 | 7 | 28 |
|  | % | 21.4 | 53.6 | 21.4 | 3.6 | 75 | 25 | 100 |
| Hypersomnolence disorders (e.g., narcolepsy, hypersomnia) | n | 7 | 15 | 2 | 2 | 22 | 4 | 26 |
|  | % | 26.9 | 57.7 | 7.7 | 7.7 | 84.6 | 15.4 | 100 |
| Insomnia | n | 1 | 5 | 17 | 9 | 6 | 26 | 32 |
|  | % | 3.1 | 15.6 | 53.1 | 28.1 | 18.7 | **81.2** | 100 |
| Sleep timing (i.e., advanced or delayed sleep phase) | n | 3 | 9 | 18 | 4 | 12 | 22 | 34 |
|  | % | 8.8 | 26.5 | 52.9 | 11.8 | 35.3 | 64.7 | 100 |
| Self-reported sleep onset latency | n | 1 | 12 | 14 | 3 | 13 | 17 | 30 |
|  | % | 3.3 | 40 | 46.7 | 10 | 43.3 | 56.7 | 100 |
| Self-reported total sleep time | n | 1 | 7 | 18 | 6 | 8 | 24 | 32 |
|  | % | 3.1 | 21.9 | 56.3 | 18.8 | 3.1 | **75.1** | 100 |
| Self-reported sleep efficiency | n | 0 | 9 | 16 | 8 | 9 | 24 | 33 |
|  | % | 0 | 27.3 | 48.5 | 24.2 | 27.3 | **72.7** | 100 |
| Self-reported nocturnal awakenings (e.g., number or duration) | n | 1 | 10 | 15 | 6 | 11 | 21 | 32 |
|  | % | 3.1 | 31.3 | 46.9 | 18.8 | 34.4 | 65.7 | 100 |
| Self-reported napping | n | 1 | 10 | 19 | 2 | 11 | 21 | 32 |
|  | % | 3.1 | 31.3 | 59.4 | 6.3 | 34.4 | 65.7 | 100 |
| **POLYSOMNOGRAPHY DATA** | | | | | | | | |
| Sleep onset latency | n | 1 | 13 | 8 | 5 | 14 | 13 | 27 |
|  | % | 3.7 | 48.1 | 29.6 | 18.5 | 51.8 | 48.1 | 100 |
| Total sleep time | n | 0 | 8 | 14 | 7 | 8 | 21 | 29 |
|  | % | 0 | 27.6 | 48.3 | 24.1 | 27.6 | **72.4** | 100 |
| Sleep efficiency | n | 0 | 7 | 14 | 8 | 7 | 22 | 29 |
|  | % | 0 | 24.1 | 48.3 | 27.6 | 24.1 | **75.9** | 100 |
| Sleep fragmentation (e.g., number of awakenings, arousals, stages shifts) | n | 0 | 6 | 15 | 11 | 6 | 26 | 32 |
|  | % | 0 | 18.8 | 46.9 | 34.4 | 18.8 | **81.3** | 100 |
| Wake after sleep onset | n | 0 | 9 | 11 | 12 | 9 | 23 | 32 |
|  | % | 0 | 28.1 | 34.4 | 37.5 | 28.1 | **71.9** | 100 |
| REM sleep latency | n | 1 | 16 | 7 | 4 | 17 | 11 | 28 |
|  | % | 3.6 | 57.1 | 25 | 14.3 | 60.7 | 39.3 | 100 |
| Duration or % of each sleep stage | n | 1 | 8 | 13 | 5 | 9 | 18 | 27 |
|  | % | 3.7 | 29.6 | 48.1 | 18.5 | 33.3 | 66.6 | 100 |
| Apnea Hypopnea Index | n | 1 | 8 | 9 | 11 | 9 | 20 | 29 |
|  | % | 3.4 | 27.6 | 31 | 37.9 | 31 | 68.9 | 100 |
| Oxygen Desaturation Index | n | 1 | 9 | 10 | 9 | 10 | 19 | 29 |
|  | % | 3.4 | 31 | 34.5 | 31 | 34.4 | 65.5 | 100 |
| NREM sleep spectral power (including slow wave activity) | n | 0 | 2 | 14 | 11 | 2 | 25 | 27 |
|  | % | 0 | 7.4 | 51.9 | 40.7 | 7.4 | **92.6** | 100 |
| REM sleep spectral power (including REM sleep EEG slowing) | n | 1 | 8 | 10 | 7 | 9 | 17 | 26 |
|  | % | 3.8 | 30.8 | 38.5 | 26.9 | 34.6 | 65.4 | 100 |
| Sleep spindles characteristics | n | 0 | 5 | 12 | 8 | 5 | 20 | 25 |
|  | % | 0 | 20 | 48 | 32 | 20 | **80** | 100 |
| Slow waves characteristics | n | 0 | 4 | 12 | 9 | 4 | 21 | 25 |
|  | % | 0 | 16 | 48 | 36 | 16 | **84** | 100 |
| K complexes | n | 0 | 10 | 6 | 3 | 10 | 9 | 19 |
|  | % | 0 | 52.6 | 31.6 | 15.8 | 52.6 | 47.4 | 100 |
| Epileptiform EEG activity | n | 3 | 11 | 2 | 3 | 14 | 5 | 19 |
|  | % | 15.8 | 57.9 | 10.5 | 15.8 | 73.7 | 26.3 | 100 |
| EEG coupling and connectivity measures | n | 0 | 4 | 12 | 4 | 4 | 16 | 20 |
|  | % | 0 | 20 | 60 | 20 | 20 | **80** | 100 |
| Features consistent with REM Sleep Behavior Disorder | n | 4 | 13 | 2 | 6 | 17 | 8 | 25 |
|  | % | 16 | 52 | 8 | 24 | 68 | 32 | 100 |
| **ACTIGRAPHY DATA** | | | | | | | | |
| Sleep onset latency | n | 0 | 14 | 9 | 6 | 14 | 15 | 29 |
|  | % | 0 | 48.3 | 31 | 20.7 | 48.3 | 51.7 | 100 |
| Total sleep time | n | 0 | 8 | 17 | 6 | 8 | 23 | 31 |
|  | % | 0 | 25.8 | 54.8 | 19.4 | 25.8 | **74.2** | 100 |
| Sleep onset and offset | n | 0 | 11 | 14 | 3 | 11 | 17 | 28 |
|  | % | 0 | 39.3 | 50 | 10.7 | 39.3 | 60.7 | 100 |
| Sleep efficiency | n | 0 | 4 | 19 | 7 | 4 | 26 | 30 |
|  | % | 0 | 13.3 | 63.3 | 23.3 | 13.3 | **86.6** | 100 |
| Wake after sleep onset | n | 0 | 4 | 18 | 8 | 4 | 26 | 30 |
|  | % | 0 | 13.3 | 60 | 26.7 | 13.3 | **86.7** | 100 |
| Daytime activity, including potential napping | n | 0 | 6 | 19 | 5 | 6 | 24 | 30 |
|  | % | 0 | 20 | 63.3 | 16.7 | 20 | **80** | 100 |
| Features derived from non-parametric or cosinor analysis | n | 0 | 6 | 13 | 6 | 6 | 19 | 25 |
|  | % | 0 | 24 | 52 | 24 | 24 | **76** | 100 |

Results of the vote are expressed in terms of frequency (n) and percentage (%). Percentages highlighted in bold reached the 70% consensus threshold. *Abbreviations: AD, Alzheimer’s disease; EEG, electroencephalography; NREM, non-rapid eye movement; OSA, obstructive sleep apnea; PLMD, periodic limb movement disorder; REM, rapid eye movement; RBD, REM sleep behavior disorder.*

**Supplementary Table 2. Sleep and circadian features sensitive to MCI – Delphi round 1.**

| **Variable** | | **Strongly Disagree** | **Disagree** | **Agree** | **Strongly Agree** | **Disagree (total)** | **Agree (total)** | **Total** |
| --- | --- | --- | --- | --- | --- | --- | --- | --- |
| **CLINICAL AND SELF-REPORTED DATA** | | | | | | | | |
| General sleep disturbance/poor sleep quality | n | 0 | 3 | 15 | 17 | 3 | 32 | 35 |
|  | % | 0 | 8.6 | 42.9 | 48.6 | 8.6 | **91.5** | 100 |
| Excessive daytime sleepiness | n | 3 | 8 | 13 | 8 | 11 | 21 | 32 |
|  | % | 9.4 | 25 | 40.6 | 25 | 34.4 | 65.6 | 100 |
| Sleep-related breathing disorders (e.g., OSA) symptoms | n | 2 | 4 | 15 | 12 | 6 | 27 | 33 |
|  | % | 6.1 | 12.1 | 45.5 | 36.4 | 18.2 | **81.9** | 100 |
| Parasomnias (e.g., RBD) and related symptoms | n | 4 | 8 | 11 | 4 | 12 | 15 | 27 |
|  | % | 14.8 | 29.6 | 40.7 | 14.8 | 44.4 | 55.5 | 100 |
| Sleep-related movement disorders (e.g., PLMD) | n | 4 | 14 | 6 | 2 | 18 | 8 | 26 |
|  | % | 15.4 | 53.8 | 23.1 | 7.7 | 69.2 | 30.8 | 100 |
| Hypersomnolence disorders (e.g., narcolepsy, hypersomnia) | n | 4 | 14 | 3 | 2 | 18 | 5 | 23 |
|  | % | 17.4 | 60.9 | 13 | 8.7 | 78.3 | 21.7 | 100 |
| Insomnia | n | 0 | 4 | 17 | 9 | 4 | 26 | 30 |
|  | % | 0 | 13.3 | 56.7 | 30 | 13.3 | **86.7** | 100 |
| Sleep timing (i.e., advanced or delayed sleep phase) | n | 1 | 8 | 12 | 8 | 9 | 20 | 29 |
|  | % | 3.4 | 27.6 | 41.4 | 27.6 | 31 | 69 | 100 |
| Self-reported sleep onset latency | n | 0 | 8 | 14 | 5 | 8 | 19 | 27 |
|  | % | 0 | 29.6 | 51.9 | 18.5 | 29.6 | **70.4** | 100 |
| Self-reported total sleep time | n | 0 | 6 | 19 | 2 | 6 | 21 | 27 |
|  | % | 0 | 22.2 | 70.4 | 7.4 | 22.2 | **77.8** | 100 |
| Self-reported sleep efficiency | n | 0 | 6 | 15 | 6 | 6 | 21 | 27 |
|  | % | 0 | 22.2 | 55.6 | 22.2 | 22.2 | **77.8** | 100 |
| Self-reported nocturnal awakenings (e.g., number or duration) | n | 1 | 7 | 13 | 6 | 8 | 19 | 27 |
|  | % | 3.7 | 25.9 | 48.1 | 22.2 | 29.6 | **70.3** | 100 |
| Self-reported napping | n | 0 | 6 | 17 | 4 | 6 | 21 | 27 |
|  | % | 0 | 22.2 | 63 | 14.8 | 22.2 | **77.8** | 100 |
| **POLYSOMNOGRAPHY DATA** | | | | | | | | |
| Sleep onset latency | n | 0 | 5 | 14 | 5 | 5 | 19 | 24 |
|  | % | 0 | 20.8 | 58.3 | 20.8 | 20.8 | **79.1** | 100 |
| Total sleep time | n | 0 | 3 | 18 | 7 | 3 | 25 | 28 |
|  | % | 0 | 10.7 | 64.3 | 25 | 10.7 | **89.3** | 100 |
| Sleep efficiency | n | 0 | 2 | 15 | 8 | 2 | 23 | 25 |
|  | % | 0 | 8 | 60 | 32 | 8 | **92** | 100 |
| Sleep fragmentation (e.g., number of awakenings, arousals, stages shifts) | n | 0 | 2 | 13 | 12 | 2 | 25 | 27 |
|  | % | 0 | 7.4 | 48.1 | 44.4 | 7.4 | **92.5** | 100 |
| Wake after sleep onset | n | 0 | 2 | 14 | 11 | 2 | 25 | 27 |
|  | % | 0 | 7.4 | 51.9 | 40.7 | 7.4 | **92.6** | 100 |
| REM sleep latency | n | 0 | 9 | 11 | 3 | 9 | 14 | 23 |
|  | % | 0 | 39.1 | 47.8 | 13 | 39.1 | 60.8 | 100 |
| Duration or % of each sleep stage | n | 0 | 4 | 15 | 6 | 4 | 21 | 25 |
|  | % | 0 | 16 | 60 | 24 | 16 | **84** | 100 |
| Apnea Hypopnea Index | n | 0 | 6 | 11 | 9 | 6 | 20 | 26 |
|  | % | 0 | 23.1 | 42.3 | 34.6 | 23.1 | **76.9** | 100 |
| Oxygen Desaturation Index | n | 0 | 6 | 11 | 9 | 6 | 20 | 26 |
|  | % | 0 | 23.1 | 42.3 | 34.6 | 23.1 | **76.9** | 100 |
| NREM sleep spectral power (including slow wave activity) | n | 0 | 1 | 13 | 12 | 1 | 25 | 26 |
|  | % | 0 | 3.8 | 50 | 46.2 | 3.8 | **96.2** | 100 |
| REM sleep spectral power (including REM sleep EEG slowing) | n | 0 | 3 | 10 | 12 | 3 | 22 | 25 |
|  | % | 0 | 12 | 40 | 48 | 12 | **88** | 100 |
| Sleep spindles characteristics | n | 0 | 2 | 11 | 9 | 2 | 20 | 22 |
|  | % | 0 | 9.1 | 50 | 40.9 | 9.1 | **90.9** | 100 |
| Slow waves characteristics | n | 0 | 3 | 11 | 10 | 3 | 21 | 24 |
|  | % | 0 | 12.5 | 45.8 | 41.7 | 12.5 | **87.5** | 100 |
| K complexes | n | 0 | 4 | 11 | 2 | 4 | 13 | 17 |
|  | % | 0 | 23.5 | 64.7 | 11.8 | 23.5 | **76.5** | 100 |
| Epileptiform EEG activity | n | 2 | 5 | 6 | 3 | 7 | 9 | 16 |
|  | % | 12.5 | 31.3 | 37.5 | 18.8 | 43.8 | 56.3 | 100 |
| EEG coupling and connectivity measures | n | 0 | 4 | 7 | 6 | 4 | 13 | 17 |
|  | % | 0 | 23.5 | 41.2 | 35.3 | 23.5 | **76.5** | 100 |
| Features consistent with REM Sleep Behavior Disorder | n | 4 | 6 | 6 | 5 | 10 | 11 | 21 |
|  | % | 19 | 28.6 | 28.6 | 23.8 | 47.6 | 52.4 | 100 |
| **ACTIGRAPHY DATA** | | | | | | | | |
| Sleep onset latency | n | 0 | 3 | 14 | 6 | 3 | 20 | 23 |
|  | % | 0 | 13 | 60.9 | 26.1 | 13 | **87** | 100 |
| Total sleep time | n | 0 | 2 | 19 | 6 | 2 | 25 | 27 |
|  | % | 0 | 7.4 | 70.4 | 22.2 | 7.4 | **92.6** | 100 |
| Sleep onset and offset | n | 0 | 6 | 13 | 5 | 6 | 18 | 24 |
|  | % | 0 | 25 | 54.2 | 20.8 | 25 | **75** | 100 |
| Sleep efficiency | n | 0 | 1 | 18 | 7 | 1 | 25 | 26 |
|  | % | 0 | 3.8 | 69.2 | 26.9 | 3.8 | **96.1** | 100 |
| Wake after sleep onset | n | 0 | 2 | 17 | 8 | 2 | 25 | 27 |
|  | % | 0 | 7.4 | 63 | 29.6 | 7.4 | **92.6** | 100 |
| Daytime activity, including potential napping | n | 0 | 1 | 17 | 8 | 1 | 25 | 26 |
|  | % | 0 | 3.8 | 65.4 | 30.8 | 3.8 | **96.2** | 100 |
| Features derived from non-parametric or cosinor analysis | n | 0 | 2 | 14 | 6 | 2 | 20 | 22 |
|  | % | 0 | 9.1 | 63.6 | 27.3 | 9.1 | **90.9** | 100 |

Results of the vote are expressed in terms of frequency (n) and percentage (%). Percentages highlighted in bold reached the 70% consensus threshold. *Abbreviations: EEG, electroencephalography; MCI, Mild Cognitive Impairment; NREM, non-rapid eye movement; OSA, obstructive sleep apnea; PLMD, periodic limb movement disorder; REM, rapid eye movement; RBD, REM sleep behavior disorder.*

**Supplementary Table 3. Sleep and circadian features sensitive to AD dementia – Delphi round 1.**

| **Variable** | | **Strongly Disagree** | **Disagree** | **Agree** | **Strongly Agree** | **Disagree (total)** | **Agree (total)** | **Total** |
| --- | --- | --- | --- | --- | --- | --- | --- | --- |
| **CLINICAL AND SELF-REPORTED DATA** | | | | | | | | |
| General sleep disturbance/poor sleep quality | n | 0 | 0 | 10 | 25 | 0 | 35 | 35 |
|  | % | 0 | 0 | 28.6 | 71.4 | 0 | **100** | 100 |
| Excessive daytime sleepiness | n | 1 | 2 | 14 | 17 | 3 | 31 | 34 |
|  | % | 2.9 | 5.9 | 41.2 | 50 | 8.8 | **91.2** | 100 |
| Sleep-related breathing disorders (e.g., OSA) symptoms | n | 2 | 2 | 14 | 13 | 4 | 27 | 31 |
|  | % | 6.5 | 6.5 | 45.2 | 41.9 | 13 | **87.1** | 100 |
| Parasomnias (e.g., RBD) and related symptoms | n | 2 | 8 | 12 | 3 | 10 | 15 | 25 |
|  | % | 8 | 32 | 48 | 12 | 40 | 60 | 100 |
| Sleep-related movement disorders (e.g., PLMD) | n | 2 | 10 | 11 | 2 | 12 | 13 | 25 |
|  | % | 8 | 40 | 44 | 8 | 48 | 52 | 100 |
| Hypersomnolence disorders (e.g., narcolepsy, hypersomnia) | n | 2 | 11 | 8 | 4 | 13 | 12 | 25 |
|  | % | 8 | 44 | 32 | 16 | 52 | 48 | 100 |
| Insomnia | n | 0 | 4 | 9 | 15 | 4 | 24 | 28 |
|  | % | 0 | 14.3 | 32.1 | 53.6 | 14.3 | **85.7** | 100 |
| Sleep timing (i.e., advanced or delayed sleep phase) | n | 0 | 1 | 11 | 19 | 1 | 30 | 31 |
|  | % | 0 | 3.2 | 35.5 | 61.3 | 3.2 | **96.8** | 100 |
| Self-reported sleep onset latency | n | 1 | 10 | 11 | 4 | 11 | 15 | 26 |
|  | % | 3.8 | 38.5 | 42.3 | 15.4 | 42.3 | 57.7 | 100 |
| Self-reported total sleep time | n | 1 | 11 | 9 | 5 | 12 | 14 | 26 |
|  | % | 3.8 | 42.3 | 34.6 | 19.2 | 46.1 | 53.8 | 100 |
| Self-reported sleep efficiency | n | 1 | 8 | 11 | 8 | 9 | 19 | 28 |
|  | % | 3.6 | 28.6 | 39.3 | 28.6 | 32.2 | 67.9 | 100 |
| Self-reported nocturnal awakenings (e.g., number or duration) | n | 1 | 8 | 11 | 8 | 9 | 19 | 28 |
|  | % | 3.6 | 28.6 | 39.3 | 28.6 | 32.2 | 67.9 | 100 |
| Self-reported napping | n | 0 | 7 | 15 | 5 | 7 | 20 | 27 |
|  | % | 0 | 25.9 | 55.6 | 18.5 | 25.9 | **74.1** | 100 |
| **POLYSOMNOGRAPHY DATA** | | | | | | | | |
| Sleep onset latency | n | 0 | 5 | 9 | 9 | 5 | 18 | 23 |
|  | % | 0 | 21.7 | 39.1 | 39.1 | 21.7 | **78.2** | 100 |
| Total sleep time | n | 0 | 3 | 13 | 12 | 3 | 25 | 28 |
|  | % | 0 | 10.7 | 46.4 | 42.9 | 10.7 | **89.3** | 100 |
| Sleep efficiency | n | 0 | 2 | 12 | 13 | 2 | 25 | 27 |
|  | % | 0 | 7.4 | 44.4 | 48.1 | 7.4 | **92.5** | 100 |
| Sleep fragmentation (e.g., number of awakenings, arousals, stages shifts) | n | 0 | 0 | 10 | 19 | 0 | 29 | 29 |
|  | % | 0 | 0 | 34.5 | 65.5 | 0 | 100 | 100 |
| Wake after sleep onset | n | 0 | 0 | 7 | 22 | 0 | 29 | 29 |
|  | % | 0 | 0 | 24.1 | 75.9 | 0 | **100** | 100 |
| REM sleep latency | n | 0 | 7 | 9 | 7 | 7 | 16 | 23 |
|  | % | 0 | 30.4 | 39.1 | 30.4 | 30.4 | 69.5 | 100 |
| Duration or % of each sleep stage | n | 0 | 1 | 12 | 13 | 1 | 25 | 26 |
|  | % | 0 | 3.8 | 46.2 | 50 | 3.8 | **96.2** | 100 |
| Apnea Hypopnea Index | n | 0 | 5 | 9 | 12 | 5 | 21 | 26 |
|  | % | 0 | 19.2 | 34.6 | 46.2 | 19.2 | **80.8** | 100 |
| Oxygen Desaturation Index | n | 0 | 5 | 8 | 12 | 5 | 20 | 25 |
|  | % | 0 | 20 | 32 | 48 | 20 | **80** | 100 |
| NREM sleep spectral power (including slow wave activity) | n | 0 | 1 | 8 | 17 | 1 | 25 | 26 |
|  | % | 0 | 3.8 | 30.8 | 65.4 | 3.8 | **96.2** | 100 |
| REM sleep spectral power (including REM sleep EEG slowing) | n | 0 | 2 | 7 | 15 | 2 | 22 | 24 |
|  | % | 0 | 8.3 | 29.2 | 62.5 | 8.3 | **91.7** | 100 |
| Sleep spindles characteristics | n | 0 | 1 | 9 | 14 | 1 | 23 | 24 |
|  | % | 0 | 4.2 | 37.5 | 58.3 | 4.2 | **95.8** | 100 |
| Slow waves characteristics | n | 0 | 2 | 7 | 17 | 2 | 24 | 26 |
|  | % | 0 | 7.7 | 26.9 | 65.4 | 7.7 | **92.3** | 100 |
| K complexes | n | 0 | 3 | 8 | 7 | 3 | 15 | 18 |
|  | % | 0 | 16.7 | 44.4 | 38.9 | 16.7 | **83.3** | 100 |
| Epileptiform EEG activity | n | 1 | 4 | 6 | 7 | 5 | 13 | 18 |
|  | % | 5.6 | 22.2 | 33.3 | 38.9 | 27.8 | **72.2** | 100 |
| EEG coupling and connectivity measures | n | 0 | 2 | 8 | 7 | 2 | 15 | 17 |
|  | % | 0 | 11.8 | 47.1 | 41.2 | 11.8 | **88.3** | 100 |
| Features consistent with REM Sleep Behavior Disorder | n | 4 | 8 | 3 | 5 | 12 | 8 | 20 |
|  | % | 20 | 40 | 15 | 25 | 60 | 40 | 100 |
| **ACTIGRAPHY DATA** | | | | | | | | |
| Sleep onset latency | n | 1 | 3 | 9 | 10 | 4 | 19 | 23 |
|  | % | 4.3 | 13 | 39.1 | 43.5 | 17.3 | **82.6** | 100 |
| Total sleep time | n | 1 | 2 | 14 | 11 | 3 | 25 | 28 |
|  | % | 3.6 | 7.1 | 50 | 39.3 | 10.7 | **89.3** | 100 |
| Sleep onset and offset | n | 0 | 4 | 11 | 11 | 4 | 22 | 26 |
|  | % | 0 | 15.4 | 42.3 | 42.3 | 15.4 | **84.6** | 100 |
| Sleep efficiency | n | 0 | 1 | 12 | 14 | 1 | 26 | 27 |
|  | % | 0 | 3.7 | 44.4 | 51.9 | 3.7 | **96.3** | 100 |
| Wake after sleep onset | n | 0 | 1 | 12 | 14 | 1 | 26 | 27 |
|  | % | 0 | 3.7 | 44.4 | 51.9 | 3.7 | **96.3** | 100 |
| Daytime activity, including potential napping | n | 0 | 1 | 13 | 14 | 1 | 27 | 28 |
|  | % | 0 | 3.6 | 46.4 | 50 | 3.6 | **96.4** | 100 |
| Features derived from non-parametric or cosinor analysis | n | 0 | 2 | 6 | 13 | 2 | 19 | 21 |
|  | % | 0 | 9.5 | 28.6 | 61.9 | 9.5 | **90.5** | 100 |

Results of the vote are expressed in terms of frequency (n) and percentage (%). Percentages highlighted in bold reached the 70% consensus threshold. *Abbreviations: AD, Alzheimer’s disease; EEG, electroencephalography; NREM, non-rapid eye movement; OSA, obstructive sleep apnea; PLMD, periodic limb movement disorder; REM, rapid eye movement; RBD, REM sleep behavior disorder.*

**Supplementary Table 4: Sleep and circadian features sensitive to pathological aging – Delphi round 2.**

| **Sleep and circadian features** | **Frequency** | **Rank** |
| --- | --- | --- |
| ***Measures sensitive to preclinical AD*** | | |
| PSG: Sleep fragmentation (e.g., number of awakenings, arousals, stages shifts) | 25 | 1 |
| Actigraphy: Sleep efficiency | 25 | 2 |
| Clinical/self-report: General sleep disturbance/poor sleep quality | 23 | 3 |
| PSG: Sleep efficiency (SE) | 23 | 4 |
| PSG: Wake after sleep onset (WASO) | 20 | 5 |
| PSG: NREM sleep spectral power (including slow wave activity) | 20 | 6 |
| Actigraphy: Wake after sleep onset (WASO) | 19 | 7 |
| Actigraphy: Total sleep time (TST) | 18 | 8 |
| PSG: Slow waves characteristics | 16 | 9 |
| Actigraphy: Daytime activity, including potential napping | 15 | 10 |
| ***Measures sensitive to MCI*** | | |
| Clinical/self-report: General sleep disturbance/poor sleep quality | 20 | 1 |
| PSG: Sleep fragmentation (e.g., number of awakenings, arousals, stages shifts) | 19 | 2 |
| Actigraphy: Sleep efficiency | 19 | 3 |
| PSG: NREM sleep spectral power (including slow wave activity) | 17 | 4 |
| Clinical/self-report: Insomnia | 15 | 5 |
| PSG: Sleep efficiency (SE) | 15 | 6 |
| PSG: Wake after sleep onset (WASO) | 15 | 7 |
| PSG: REM sleep spectral power (including REM sleep EEG slowing) | 15 | 8 |
| PSG: Duration in each sleep stage/% | 14 | 9 |
| Actigraphy: Wake after sleep onset (WASO) | 14 | 10 |
| ***Measures sensitive to AD dementia*** | | |
| PSG: Sleep fragmentation (e.g., number of awakenings, arousals, stages shifts) | 20 | 1 |
| PSG: NREM sleep spectral power (including slow wave activity) | 19 | 2 |
| PSG: REM sleep spectral power (including REM sleep EEG slowing) | 17 | 3 |
| PSG: Slow waves characteristics | 16 | 4 |
| Actigraphy: Wake after sleep onset (WASO) | 16 | 5 |
| Clinical/self-report: General sleep disturbance/poor sleep quality | 15 | 6 |
| Clinical/self-report: Excessive daytime sleepiness | 15 | 7 |
| PSG: Sleep efficiency (SE) | 15 | 8 |
| PSG: Wake after sleep onset (WASO) | 15 | 9 |
| Actigraphy: Daytime activity, including potential napping | 15 | 10 |

Results of the vote are expressed in terms of frequencies and ranks, determined by a sum of frequencies approach. For clarity, only the 10 best ranking items in each category are reported in the table. *Abbreviations: AD, Alzheimer’s disease; EEG, electroencephalography; MCI, Mild Cognitive Impairment; NREM, non-rapid eye movement; PSG, polysomnography.*

**Supplementary Table 5. Best method to assess sleep and circadian features.**

| **DELPHI ROUND 1** | | | | | | | |
| --- | --- | --- | --- | --- | --- | --- | --- |
| **Variable** | **Method** | **Preclinical AD** | | **MCI** | | **AD dementia** | |
|  |  | n | % | n | % | n | % |
| Time in Bed (TIB) | Patient self-report | 10 | 27 | 3 | 8.3 | 0 | 0 |
|  | Informant report | 0 | 0 | 3 | 8.3 | 9 | 25.7 |
|  | Accelerometer/wearable devices | 17 | 45.9 | 15 | 41.7 | 15 | 42.9 |
|  | In-lab PSG/EEG | 6 | 16.2 | 7 | 19.4 | 5 | 14.3 |
|  | At-home PSG/EEG | 4 | 10.8 | 8 | 22.2 | 6 | 17.1 |
|  | Total | 37 | 100 | 36 | 100 | 35 | 100 |
| Sleep duration/total sleep time (TST) | Patient self-report | 1 | 2.6 | 0 | 0 | 0 | 0 |
|  | Informant report | 0 | 0 | 0 | 0 | 1 | 2.9 |
|  | Accelerometer/wearable devices | 13 | 34.2 | 8 | 22.2 | 12 | 34.3 |
|  | In-lab PSG/EEG | 9 | 23.7 | 10 | 27.8 | 6 | 17.1 |
|  | At-home PSG/EEG | 15 | 39.5 | 18 | 50 | 16 | 45.7 |
|  | Total | 38 | 100 | 36 | 100 | 35 | 100 |
| Sleep onset latency (SOL) | Patient self-report | 3 | 8.1 | 1 | 2.7 | 0 | 0 |
|  | Informant report | 0 | 0 | 0 | 0 | 2 | 5.6 |
|  | Accelerometer/wearable devices | 7 | 18.9 | 5 | 13.5 | 7 | 19.4 |
|  | In-lab PSG/EEG | 12 | 32.4 | 13 | 35.1 | 9 | 25 |
|  | At-home PSG/EEG | 15 | 40.5 | 18 | 48.6 | 18 | 50 |
|  | Total | 37 | 100 | 37 | 100 | 36 | 100 |
| Sleep efficiency | Patient self-report | 1 | 2.6 | 0 | 0 | 0 | 0 |
|  | Informant report | 0 | 0 | 0 | 0 | 1 | 2.9 |
|  | Accelerometer/wearable devices | 11 | 28.9 | 8 | 21.6 | 10 | 28.6 |
|  | In-lab PSG/EEG | 9 | 23.7 | 12 | 32.4 | 6 | 17.1 |
|  | At-home PSG/EEG | 17 | 44.7 | 17 | 45.9 | 18 | 51.4 |
|  | Total | 38 | 100 | 37 | 100 | 35 | 100 |
| Sleep fragmentation (e.g., awakenings) | Patient self-report | 1 | 2.7 | 0 | 0 | 0 | 0 |
|  | Informant report | 0 | 0 | 0 | 0 | 2 | 5.6 |
|  | Accelerometer/wearable devices | 8 | 21.6 | 6 | 16.2 | 9 | 25 |
|  | In-lab PSG/EEG | 14 | 37.8 | 16 | 43.2 | 8 | 22.2 |
|  | At-home PSG/EEG | 14 | 37.8 | 15 | 40.5 | 17 | 47.2 |
|  | Total | 37 | 100 | 37 | 100 | 36 | 100 |
| Wake after sleep onset (WASO) | Patient self-report | 1 | 2.6 | 0 | 0 | 0 | 0 |
|  | Informant report | 0 | 0 | 0 | 0 | 1 | 2.8 |
|  | Accelerometer/wearable devices | 8 | 21.1 | 5 | 14.3 | 9 | 25 |
|  | In-lab PSG/EEG | 14 | 36.8 | 14 | 40 | 8 | 22.2 |
|  | At-home PSG/EEG | 15 | 39.5 | 16 | 45.7 | 18 | 50 |
|  | Total | 38 | 100 | 35 | 100 | 36 | 100 |
| Naps (timing. duration. frequency) | Patient self-report | 9 | 23.7 | 2 | 5.6 | 0 | 0 |
|  | Informant report | 3 | 7.9 | 8 | 22.2 | 12 | 34.3 |
|  | Accelerometer/wearable devices | 21 | 55.3 | 20 | 55.6 | 19 | 54.3 |
|  | In-lab PSG/EEG | 3 | 7.9 | 4 | 11.1 | 2 | 5.7 |
|  | At-home PSG/EEG | 2 | 5.3 | 2 | 5.6 | 2 | 5.7 |
|  | Total | 38 | 100 | 36 | 100 | 35 | 100 |
| Sleep-disordered breathing | Patient self-report | 1 | 2.6 | 1 | 2.7 | 0 | 0 |
|  | Informant report | 0 | 0 | 0 | 0 | 0 | 0 |
|  | At home oximetry monitors | 2 | 5.3 | 2 | 5.4 | 4 | 10.8 |
|  | In-lab PSG/sleep apnea test | 15 | 39.5 | 14 | 37.8 | 11 | 29.7 |
|  | At-home PSG/ sleep apnea test | 20 | 52.6 | 20 | 54.1 | 22 | 59.5 |
|  | Total | 38 | 100 | 37 | 100 | 37 | 100 |
| **DELPHI ROUND 2** | | | | | | | |
| **Measure** | **Method** | **Preclinical AD** | | **MCI** | | **AD dementia** | |
|  |  | **n** | **%** | **n** | **%** | **n** | **%** |
| Time in bed | Informant self-report | 2 | 6.5 | 7 | 22.6 | 11 | 35.5 |
|  | Patient self-report | 11 | 35.5 | 3 | 9.7 | 1 | 3.2 |
|  | Actigraphy | 12 | 38.7 | 14 | 45.2 | 12 | 38.7 |
|  | PSG/EEG | 6 | 19.4 | 7 | 22.6 | 7 | 22.6 |
|  | *Total* | *31* | *100.0* | *31* | *100.0* | *31* | *100.0* |
| Total sleep time | Informant self-report | 0 | 0.0 | 0 | 0.0 | 1 | 3.2 |
|  | Patient self-report | 2 | 6.5 | 1 | 3.2 | 0 | 0.0 |
|  | Actigraphy | 13 | 41.9 | 11 | 35.5 | 15 | 48.4 |
|  | PSG/EEG | 16 | 51.6 | 19 | 61.3 | 15 | 48.4 |
|  | *Total* | *31* | *100.0* | *31* | *100.0* | *31* | *100.0* |
| Sleep onset latency | Informant self-report | 1 | 3.3 | 0 | 0.0 | 1 | 3.3 |
|  | Patient self-report | 2 | 6.7 | 2 | 6.7 | 0 | 0.0 |
|  | Actigraphy | 5 | 16.7 | 4 | 13.3 | 10 | 33.3 |
|  | PSG/EEG | 22 | **73.3** | 24 | **80.0** | 19 | 63.3 |
|  | *Total* | *30* | *100.0* | *30* | *100.0* | *30* | *100.0* |
| Sleep efficiency | Informant self-report | 0 | 0.0 | 0 | 0.0 | 0 | 0.0 |
|  | Patient self-report | 1 | 3.2 | 0 | 0.0 | 0 | 0.0 |
|  | Actigraphy | 11 | 35.5 | 12 | 38.7 | 14 | 45.2 |
|  | PSG/EEG | 19 | 61.3 | 19 | 61.3 | 17 | 54.8 |
|  | *Total* | *31* | *100.0* | *31* | *100.0* | *31* | *100.0* |
| Sleep fragmentation (e.g., awakenings) | Informant self-report | 0 | 0.0 | 0 | 0.0 | 0 | 0.0 |
|  | Patient self-report | 1 | 3.3 | 0 | 0.0 | 0 | 0.0 |
|  | Actigraphy | 6 | 20.0 | 6 | 20.0 | 10 | 33.3 |
|  | PSG/EEG | 23 | **76.7** | 24 | **80.0** | 20 | 66.7 |
|  | *Total* | *30* | *100.0* | *30* | *100.0* | *30* | *100.0* |
| Wake after sleep onset | Informant self-report | 0 | 0.0 | 0 | 0.0 | 0 | 0.0 |
|  | Patient self-report | 0 | 0.0 | 0 | 0.0 | 0 | 0.0 |
|  | Actigraphy | 9 | 30.0 | 8 | 25.0 | 13 | 43.3 |
|  | PSG/EEG | 21 | **70.0** | 24 | **75.0** | 17 | 56.7 |
|  | *Total* | *30* | *100.0* | *32* | *100.0* | *30* | *100.0* |
| Naps (e.g., timing, duration, frequency) | Informant self-report | 2 | 6.7 | 5 | 17.2 | 9 | 30.0 |
|  | Patient self-report | 6 | 20.0 | 2 | 6.9 | 0 | 0.0 |
|  | Actigraphy | 20 | 66.7 | 21 | **72.4** | 20 | 66.7 |
|  | PSG/EEG | 2 | 6.7 | 1 | 3.4 | 1 | 3.3 |
|  | *Total* | *30* | *100.0* | *29* | *100.0* | *30* | *100.0* |

Results of the vote are expressed in terms of frequency (n) and percentage (%). Percentages highlighted in bold reached the 70% consensus threshold. *Abbreviations: AD, Alzheimer’s disease; EEG, electroencephalography; MCI, Mild Cognitive Impairment; PSG, polysomnography.*

**Supplementary Table 6. Minimum standard for data collection – Delphi round 1.**

| **Variable** |  | **Absolutely essential** | **Necessary for a well-defined study** | **Not essential- optional** | **Agreed (total)** | **Total** |
| --- | --- | --- | --- | --- | --- | --- |
| **Demographics, clinical and biological features** | | | | | | |
| Gender | n | 23 | 9 | 4 | 32 | 36 |
|  | % | 63.9 | 25 | 11.1 | **88.9** | 100 |
| Cognitive diagnosis | n | 33 | 4 | 0 | 37 | 37 |
|  | % | 89.2 | 10.8 | 0 | **100** | 100 |
| Subjective cognitive/memory concerns | n | 9 | 23 | 6 | 32 | 38 |
|  | % | 23.7 | 60.5 | 15.8 | **84.2** | 100 |
| Dementia biomarker profile for clinical staging or A/T/N classification | n | 11 | 25 | 2 | 36 | 38 |
|  | % | 28.9 | 65.8 | 5.3 | **94.7** | 100 |
| Race | n | 12 | 17 | 8 | 29 | 37 |
|  | % | 32.4 | 45.9 | 21.6 | **78.3** | 100 |
| Ethnicity | n | 9 | 19 | 9 | 28 | 37 |
|  | % | 24.3 | 51.4 | 24.3 | **75.7** | 100 |
| Education level | n | 21 | 16 | 1 | 37 | 38 |
|  | % | 55.3 | 42.1 | 2.6 | **97.4** | 100 |
| Living arrangement/residential setting | n | 6 | 20 | 11 | 26 | 37 |
|  | % | 16.2 | 54.1 | 29.7 | **70.3** | 100 |
| Social network | n | 3 | 9 | 24 | 12 | 36 |
|  | % | 8.3 | 25 | 66.7 | 33.3 | 100 |
| Employment | n | 5 | 18 | 13 | 23 | 36 |
|  | % | 13.9 | 50 | 36.1 | 63.9 | 100 |
| Vocational status (e.g., retirement) | n | 4 | 20 | 12 | 24 | 36 |
|  | % | 11.1 | 55.6 | 33.3 | 66.7 | 100 |
| Caring responsibilities | n | 3 | 19 | 14 | 22 | 36 |
|  | % | 8.3 | 52.8 | 38.9 | 61.1 | 100 |
| APOE ε4 | n | 13 | 19 | 5 | 32 | 37 |
|  | % | 35.1 | 51.4 | 13.5 | **86.5** | 100 |
| Body mass index | n | 20 | 12 | 5 | 32 | 37 |
|  | % | 54.1 | 32.4 | 13.5 | **86.5** | 100 |
| Blood pressure | n | 12 | 12 | 13 | 24 | 37 |
|  | % | 32.4 | 32.4 | 35.1 | 64.8 | 100 |
| Lifestyle: physical activity levels | n | 8 | 19 | 10 | 27 | 37 |
|  | % | 21.6 | 51.4 | 27 | **73** | 100 |
| Lifestyle: diet | n | 4 | 17 | 16 | 21 | 37 |
|  | % | 10.8 | 45.9 | 43.2 | 56.7 | 100 |
| Coffee, tea, energetic drinks consumption | n | 9 | 18 | 10 | 27 | 37 |
|  | % | 24.3 | 48.6 | 27 | **72.9** | 100 |
| Daily light exposure | n | 8 | 19 | 10 | 27 | 37 |
|  | % | 21.6 | 51.4 | 27 | **73** | 100 |
| Season (of data collection/clinical assessment) | n | 5 | 19 | 11 | 24 | 35 |
|  | % | 14.3 | 54.3 | 31.4 | 68.6 | 100 |
| Sleeping arrangement | n | 8 | 18 | 11 | 26 | 37 |
|  | % | 21.6 | 48.6 | 29.7 | **70.2** | 100 |
| Number of pregnancies | n | 0 | 11 | 25 | 11 | 36 |
|  | % | 0 | 30.6 | 69.4 | 30.6 | 100 |
| Menopause: current status | n | 6 | 20 | 10 | 26 | 36 |
|  | % | 16.7 | 55.6 | 27.8 | **72.3** | 100 |
| Menopause: age of onset | n | 3 | 19 | 14 | 22 | 36 |
|  | % | 8.3 | 52.8 | 38.9 | 61.1 | 100 |
| **Comorbidities (current and/or history)** | | | | | | |
| Smoking | n | 12 | 19 | 7 | 31 | 38 |
|  | % | 31.6 | 50 | 18.4 | **81.6** | 100 |
| Alcohol intake | n | 15 | 20 | 3 | 35 | 38 |
|  | % | 39.5 | 52.6 | 7.9 | **92.1** | 100 |
| Cannabis/Tetrahydrocannabinol (THC) | n | 13 | 15 | 9 | 28 | 37 |
|  | % | 35.1 | 40.5 | 24.3 | **75.6** | 100 |
| Illicit substances | n | 13 | 15 | 9 | 28 | 37 |
|  | % | 35.1 | 40.5 | 24.3 | **75.6** | 100 |
| State anxiety | n | 14 | 17 | 7 | 31 | 38 |
|  | % | 36.8 | 44.7 | 18.4 | **81.5** | 100 |
| Trait anxiety | n | 11 | 21 | 6 | 32 | 38 |
|  | % | 28.9 | 55.3 | 15.8 | **84.2** | 100 |
| Depressive symptoms | n | 23 | 15 | 0 | 38 | 38 |
|  | % | 60.5 | 39.5 | 0 | **100** | 100 |
| Cardiovascular diseases | n | 13 | 21 | 3 | 34 | 37 |
|  | % | 35.1 | 56.8 | 8.1 | **91.9** | 100 |
| Neurological diseases (e.g., stroke, epilepsy, Parkinson’s disease) | n | 28 | 9 | 0 | 37 | 37 |
|  | % | 75.7 | 24.3 | 0 | **100** | 100 |
| Psychiatric disorders and history of mental illness | n | 29 | 7 | 1 | 36 | 37 |
|  | % | 78.4 | 18.9 | 2.7 | **97.3** | 100 |
| Metabolic syndromes/disorders | n | 15 | 16 | 6 | 31 | 37 |
|  | % | 40.5 | 43.2 | 16.2 | **83.7** | 100 |
| Cancer | n | 11 | 14 | 11 | 25 | 36 |
|  | % | 30.6 | 38.9 | 30.6 | 69.5 | 100 |
| Traumatic brain injury | n | 18 | 17 | 2 | 35 | 37 |
|  | % | 48.6 | 45.9 | 5.4 | **94.5** | 100 |
| **Sleep** | | | | | | |
| Chronotype | n | 5 | 24 | 6 | 29 | 35 |
|  | % | 14.3 | 68.6 | 17.1 | **82.9** | 100 |
| History of shift work | n | 7 | 20 | 9 | 27 | 36 |
|  | % | 19.4 | 55.6 | 25 | **75** | 100 |
| Current shift work | n | 20 | 14 | 3 | 34 | 37 |
|  | % | 54.1 | 37.8 | 8.1 | **91.9** | 100 |
| Obstructive sleep apnea diagnosis | n | 24 | 14 | 0 | 38 | 38 |
|  | % | 63.2 | 36.8 | 0 | **100** | 100 |
| Obstructive sleep apnea treatment (e.g., continuous positive airway pressure use) | n | 25 | 12 | 0 | 37 | 37 |
|  | % | 67.6 | 32.4 | 0 | **100** | 100 |
| Insomnia symptoms | n | 20 | 14 | 3 | 34 | 37 |
|  | % | 54.1 | 37.8 | 8.1 | **91.9** | 100 |
| Insomnia treatment (e.g., CBT-I) | n | 17 | 14 | 5 | 31 | 36 |
|  | % | 47.2 | 38.9 | 13.9 | **86.1** | 100 |
| Restless legs syndrome | n | 12 | 14 | 8 | 26 | 34 |
|  | % | 35.3 | 41.2 | 23.5 | **76.5** | 100 |
| Heart rate variability | n | 4 | 11 | 21 | 15 | 36 |
|  | % | 11.1 | 30.6 | 58.3 | 41.7 | 100 |
| Others (open field): RBD and other parasomnias | n | - | - | - | 8 | - |
| Others (open field): Circadian rhythm disorders | n | - | - | - | 2 | - |
| **Medication** | | | | | | |
| Benzodiazepines | n | 26 | 11 | 0 | 37 | 37 |
|  | % | 70.3 | 29.7 | 0 | **100** | 100 |
| Non-benzodiazepine receptor agonist hypnotics | n | 24 | 12 | 0 | 36 | 36 |
|  | % | 66.7 | 33.3 | 0 | **100** | 100 |
| Trazadone | n | 22 | 11 | 1 | 33 | 34 |
|  | % | 64.7 | 32.4 | 2.9 | **97.1** | 100 |
| Other approved hypnotics (e.g., DORAs, ramelteon…) | n | 24 | 10 | 2 | 34 | 36 |
|  | % | 66.7 | 27.8 | 5.6 | **94.5** | 100 |
| Antidepressants | n | 25 | 11 | 0 | 36 | 36 |
|  | % | 69.4 | 30.6 | 0 | **100** | 100 |
| Antipsychotics | n | 23 | 11 | 2 | 34 | 36 |
|  | % | 63.9 | 30.6 | 5.6 | **94.5** | 100 |
| Opioids | n | 21 | 11 | 4 | 32 | 36 |
|  | % | 58.3 | 30.6 | 11.1 | **88.9** | 100 |
| Antihistamines | n | 15 | 13 | 8 | 28 | 36 |
|  | % | 41.7 | 36.1 | 22.2 | **77.8** | 100 |
| Acetylcholinesterase (ACE) inhibitors | n | 21 | 10 | 4 | 31 | 35 |
|  | % | 60 | 28.6 | 11.4 | **88.6** | 100 |
| Memantine | n | 20 | 11 | 3 | 31 | 34 |
|  | % | 58.8 | 32.4 | 8.8 | **91.2** | 100 |
| Monoclonal antibody | n | 11 | 16 | 3 | 27 | 30 |
|  | % | 36.7 | 53.3 | 10 | **90** | 100 |
| Non-narcotic analgesics | n | 7 | 17 | 9 | 24 | 33 |
|  | % | 21.2 | 51.5 | 27.3 | **72.7** | 100 |
| Narcotic analgesics | n | 16 | 13 | 5 | 29 | 34 |
|  | % | 47.1 | 38.2 | 14.7 | **85.3** | 100 |
| Anticonvulsants | n | 14 | 15 | 4 | 29 | 33 |
|  | % | 42.4 | 45.5 | 12.1 | **87.9** | 100 |
| Stimulants (e.g., Ritalin) | n | 18 | 13 | 2 | 31 | 33 |
|  | % | 54.5 | 39.4 | 6.1 | **93.9** | 100 |
| Hormone therapy | n | 7 | 20 | 7 | 27 | 34 |
|  | % | 20.6 | 58.8 | 20.6 | **79.4** | 100 |
| Melatonin | n | 22 | 14 | 1 | 36 | 37 |
|  | % | 59.5 | 37.8 | 2.7 | **97.3** | 100 |
| Vitamins and supplements | n | 2 | 14 | 20 | 16 | 36 |
|  | % | 5.6 | 38.9 | 55.6 | 44.5 | 100 |
| Phytotherapy and homeopathy | n | 1 | 10 | 24 | 11 | 35 |
|  | % | 2.9 | 28.6 | 68.6 | 31.5 | 100 |

Results of the vote are expressed in terms of frequency (n) and percentage (%). Percentages highlighted in bold reached the 70% consensus threshold. *Abbreviations: A/T/N, amyloid/tau/neurodegeneration; APOE, apolipoprotein E; CBT-I, Cognitive behavioral therapy for insomnia; DORAs, Dual Orexin Receptor Antagonists; RBD, REM sleep behavior disorder.*

**Supplementary Table 7. Minimum standard for data collection – Delphi round 2.**

| **Feature** | **n/32** | **Rank** |
| --- | --- | --- |
| **Demographic, clinical, and biological features** | | |
| Cognitive diagnosis | 28 | **1** |
| Biomarker profile for clinical staging or A/T/N classification | 24 | **2** |
| *APOE ε4* | 21 | **3** |
| Education level | 17 | **4** |
| Body mass index | 14 | **5** |
| Gender | 12 | 6 |
| Subjective cognitive/memory concerns | 7 | 7 |
| Lifestyle: physical activity levels | 7 | 7 |
| Menopause: current status | 7 | 7 |
| Sleeping arrangement | 6 | 8 |
| Ethnicity | 5 | 9 |
| Living arrangement/ residential setting | 5 | 9 |
| Race | 4 | 10 |
| Coffee, tea, energetic drinks consumption | 4 | 10 |
| Daily light exposure | 2 | 11 |
| **Comorbidities (current and/or history)** | | |
| Neurological diseases (e.g., stroke, epilepsy, Parkinson’s disease) | 29 | **1** |
| Depressive symptoms | 27 | **2** |
| Psychiatric disorders and history of mental illness | 24 | **3** |
| Cardiovascular diseases | 17 | **4** |
| Alcohol intake | 16 | **5** |
| Smoking | 11 | 6 |
| Metabolic syndromes/disorders | 11 | 6 |
| State anxiety | 10 | 7 |
| Traumatic brain injury | 7 | 8 |
| Illicit substances | 4 | 9 |
| Trait anxiety | 4 | 9 |
| Cannabis/Tetrahydrocannabinol (THC) | 3 | 10 |
| **Sleep-related features** | | |
| Insomnia symptoms | 30 | **1** |
| OSA diagnosis | 29 | **2** |
| OSA treatment (e.g., continuous positive airway pressure use) | 26 | **3** |
| Current shift work | 22 | **4** |
| Restless legs syndrome | 16 | **5** |
| Chronotype | 13 | 6 |
| Insomnia treatment (e.g., CBT-I) | 13 | 6 |
| History of shift work | 6 | 7 |
| **Medication** | | |
| Benzodiazepines | 28 | **1** |
| Non-benzodiazepine receptor agonist hypnotics | 22 | **2** |
| Antidepressants | 20 | **3** |
| Other approved hypnotics (e.g., DORAs, ramelteon) | 17 | **4** |
| Trazodone | 12 | **5** |
| Antipsychotics | 12 | **5** |
| Opioids | 7 | 6 |
| Acetylcholinesterase (ACE) inhibitors | 7 | 6 |
| Melatonin | 7 | 6 |
| Stimulants (e.g., Ritalin) | 6 | 7 |
| Anticonvulsants | 4 | 8 |
| Memantine | 3 | 9 |
| Antihistamines | 2 | 10 |
| Monoclonal antibody | 2 | 10 |
| Narcotic analgesics | 2 | 10 |
| Hormone therapy | 2 | 10 |
| Non-narcotic analgesics | 0 | 11 |

Results of the vote are expressed in terms of frequency (n/32) and rank, determine using a sum of frequencies approach. The 5 best-ranking options in each category are highlighted in bold. *Abbreviations: A/T/N, amyloid/tau/neurodegeneration; APOE, apolipoprotein E; CBT-I, Cognitive behavioral therapy for insomnia; DORAs, Dual Orexin Receptor Antagonists; OSA, obstructive sleep apnea.*

**Supplementary Table 8. Confidence in self-reported sleep data.**

| **Question** | **Answer options** | **n** | **%** |
| --- | --- | --- | --- |
| How confident are you in the reliability of single-item subjective self- or informant-reported sleep measures? | Not confident at all | 10 | 26.3 |
|  | Not confident | 19 | 50 |
|  | Confident | 7 | 18.4 |
|  | Very confident | 2 | 5.3 |
|  | *Not confident (total)* | *29* | ***76.3*** |
|  | *Confident (total)* | *9* | *23.7* |
|  | *Total* | *38* | *100* |
| Do you feel that the quality of self-reported data depends on the cognitive status of the patient/participant? | Yes | 38 | **100** |
|  | No | 0 | 0 |
|  | *Total* | *38* | *100* |
| How confident are you in informant-related sleep data? | Not confident at all | 3 | 7.9 |
|  | Not confident | 20 | 52.6 |
|  | Confident | 13 | 34.2 |
|  | Very confident | 2 | 5.3 |
|  | *Not confident (total)* | *23* | *60.5* |
|  | *Confident (total)* | *15* | *39.5* |
|  | *Total* | *38* | *100* |
| Do you feel the reliability of informant-related sleep data depends on the cognitive status of the informant? | Yes | 37 | **97.4** |
|  | No | 1 | 2.6 |
|  | *Total* | *38* | *100* |
| Do you feel that sleep questionnaires should be re-validated for the ageing/MCI/AD population? | Yes | 35 | **92.1** |
|  | No | 3 | 7.9 |
|  | *Total* | *38* | *100* |
| Do you feel that the scoring of sleep questionnaires should be adapted/standardized based on the ageing, MCI or AD population? | Yes | 36 | **94.7** |
|  | No | 2 | 5.3 |
|  | *Total* | *38* | *100* |

Results of the vote are expressed in terms of frequency (n) and percentage (%). Percentages highlighted in bold reached the 70% consensus threshold. *Abbreviations: AD, Alzheimer’s disease; MCI, Mild Cognitive Impairment.*

**Supplementary Table 9. Confidence and recommendations for actigraphy data.**

| **Question** | **Answer options** | **n** | **%** |
| --- | --- | --- | --- |
| In a research context, should a sleep diary always be proposed in conjunction with actigraphy? | Yes | 33 | **86.8** |
|  | No | 5 | 13.2 |
|  | *Total* | *38* | *100* |
| What questions should be asked in a sleep diary? | Sleep latency | 32 | - |
|  | Use of caffeine | 34 | - |
|  | Use of alcohol | 34 | - |
|  | Type of sleep disruptions | 24 | - |
|  | Wake time | 35 | - |
|  | Feeling refreshed upon waking | 32 | - |
|  | Other: sleep timing (n=9), daytime napping (n=5), presence of a bed partner (n=1), light exposure (n=1), sleep medication (n=1), usual pattern of sleep (n=1), reason for sleep disruption (n=1), snoring (n=1), gasp for air (n=1), morning headaches (n=1), restless legs (n=1). | | |
| How confident are you in sleep diary data as a tool for quality control? How much do you rely on it to interpret your recordings? | Not confident | 12 | 33.3 |
|  | Confident | 22 | 61.1 |
|  | Very confident | 2 | 5.6 |
|  | *Not confident (total)* | *12* | *33.3* |
|  | *Confident (total)* | *24* | *66.7* |
|  | *Total* | *36* | *100* |
| How many days of actigraphy recordings are the minimum standard to measure sleep in a research setting? | <7 days | 6 | 19.4 |
|  | 7 to 13 days | 22 | **71** |
|  | ≥ 14 days | 3 | 9.7 |
|  | *Total* | *31* | *100* |
| How many days of actigraphy recordings are ideal to measure sleep in a research setting? | <7 days | 1 | 3.2 |
|  | 7 to 13 days | 9 | 29 |
|  | ≥ 14 days | 21 | 67.7 |
|  | *Total* | *31* | *100* |
| How many days of actigraphy recordings are the minimum standard to measure rest-activity in a research setting? | <7 days | 4 | 12.9 |
|  | 7 to 13 days | 19 | 61.3 |
|  | ≥ 14 days | 8 | 25.8 |
|  | *Total* | *31* | *100* |
| How many days of actigraphy recordings are ideal to measure rest-activity in a research setting? | <7 days | 0 | 0 |
|  | 7 to 13 days | 7 | 22.6 |
|  | ≥ 14 days | 24 | **77.4** |
|  | *Total* | *31* | *100* |
| Do you alter the actigraphy recording based on diary report? | Yes | 11 | 32.4 |
|  | No | 2 | 5.9 |
|  | Sometimes | 21 | 61.8 |
|  | *Total* | *34* | *100* |
| Would you recommend using software-implemented algorithms or in-house processing for actigraphy? | Software-implemented | 1 | 3.6 |
|  | In-house | 1 | 3.6 |
|  | Both in-house and software-implemented | 26 | **92.9** |
|  | *Total* | *28* | *100* |

Results of the vote are expressed in terms of frequency (n) and percentage (%). Percentages highlighted in bold reached the 70% consensus threshold.

**Supplementary Table 10. Recommendations for polysomnography and EEG data.**

| **Question** | **Answer options** | **n** | **%** |
| --- | --- | --- | --- |
| What minimum EEG montage do you recommend in a research setting? | Fp1 | 9 | 22.5 |
|  | Fp2 | 9 | 22.5 |
|  | F3 | 18 | 45 |
|  | F4 | 19 | 47.5 |
|  | F7 | 4 | 10 |
|  | F8 | 4 | 10 |
|  | Fz | 12 | 30 |
|  | T3 | 7 | 17.5 |
|  | T4 | 7 | 17.5 |
|  | T5 | 6 | 15 |
|  | T6 | 6 | 15 |
|  | C3 | 16 | 40 |
|  | C4 | 17 | 42.5 |
|  | Cz | 15 | 37.5 |
|  | P3 | 9 | 22.5 |
|  | P4 | 9 | 22.5 |
|  | Pz | 11 | 27.5 |
|  | O1 | 18 | 45 |
|  | O2 | 19 | 47.5 |
|  | Comment: *P9, P10, T9, T10, F9, F10 (n=1)* | | |
| For cohort studies, do you feel a habituation night should be systematically performed? | No | 14 | 40 |
|  | Yes, always (regardless of the clinical group) | 20 | 57.1 |
|  | Only in cognitively unimpaired participants, but not MCI/dementia patients | 1 | 2.9 |
|  | *Total* | *35* | *100* |
| For optimal compromise between feasibility and ecological validity, would you favor in-home or in-lab settings for future protocols and clinical trials? | Always in-lab PSG regardless of the clinical group and research question | 1 | 2.7 |
|  | Always at-home PSG regardless of the clinical group and research question | 2 | 5.4 |
|  | The choice depends on the research question only | 1 | 2.7 |
|  | The choice depends on the clinical group only | 1 | 2.7 |
|  | The choice depends on both the question and the clinical group | 32 | **86.5** |
|  | *Total* | *37* | *100* |

Results of the vote are expressed in terms of frequency (n) and percentage (%). Percentages highlighted in bold reached the 70% consensus threshold. *Abbreviations: EEG, electroencephalography; MCI, Mild Cognitive Impairment; PSG, polysomnography.*

**Supplementary Table 11. Preference for in-lab or at-home PSG/EEG settings.**

| **Preferred setting** | **Preclinical AD** | | **MCI** | | **AD Dementia** | |
| --- | --- | --- | --- | --- | --- | --- |
|  | **n** | **%** | **n** | **%** | **n** | **%** |
| In-lab preferred | 8 | 22.9 | 9 | 27.3 | 9 | 27.3 |
| At-home preferred | 10 | 28.6 | 9 | 27.3 | 19 | 57.6 |
| Both are equally acceptable | 17 | 48.6 | 15 | 45.5 | 5 | 15.2 |
| *Total* | *35* | *100* | *33* | *100* | *33* | *100* |

Results of the vote are expressed in terms of frequency (n) and percentage (%). *Abbreviations: AD, Alzheimer’s disease; EEG, electroencephalography; MCI, Mild Cognitive Impairment; PSG, polysomnography.*

**Supplementary Table 12. Confidence in the use of newly available wearable and nearable devices.**

| **Question** | **Answer options** | **n** | **%** |
| --- | --- | --- | --- |
| **Accelerometer devices** | | | |
| How confident are you in movement-based devices recording movement only? | Not confident | 9 | 26.5 |
|  | Confident | 21 | 61.8 |
|  | Very confident | 4 | 11.8 |
|  | *Confident (total)* | *25* | ***73.5*** |
|  | *Total* | *34* | *100* |
| How confident are you in the use of movement-based devices combining movement recordings with other types of data (e.g., heart rate, O2). | Not confident | 7 | 21.2 |
|  | Confident | 21 | 63.6 |
|  | Very confident | 5 | 15.2 |
|  | *Confident (total)* | *26* | ***78.8*** |
|  | *Total* | *33* | *100* |
| How confident are you in the validity of “research-quality” devices in cognitively unimpaired older adults? (e.g., Actigraph, GENEActiv, etc) | Not confident | 2 | 5.6 |
|  | Confident | 25 | 69.4 |
|  | Very confident | 9 | 25 |
|  | *Confident (total)* | *34* | ***94.4*** |
|  | *Total* | *36* | *100* |
| How confident are you in the validity of “commercially-available” devices in cognitively unimpaired older adults? (e.g., Fitbit, Apple Watch, etc) | Not confident at all | 9 | 25.7 |
|  | Not confident | 16 | 45.7 |
|  | Confident | 8 | 22.9 |
|  | Very confident | 2 | 5.7 |
|  | *Not confident (total)* | *25* | ***71.4*** |
|  | *Confident (total)* | *10* | *28.6* |
|  | *Total* | *35* | *100* |
| How confident are you in the validity of “research-quality” devices in MCI/dementia patients? (e.g., Actigraph, GENEActiv, etc) | Not confident at all | 1 | 2.9 |
|  | Not confident | 6 | 17.1 |
|  | Confident | 23 | 65.7 |
|  | Very confident | 5 | 14.3 |
|  | *Not confident (total)* | 7 | 20 |
|  | *Confident (total)* | 28 | **80** |
|  | *Total* | *35* | *100* |
| How confident are you in the validity of “commercially-available” devices in MCI/dementia patients? (e.g., Fitbit, Apple Watch, etc) | Not confident at all | 10 | 30.3 |
|  | Not confident | 17 | 51.5 |
|  | Confident | 5 | 15.2 |
|  | Very confident | 1 | 3 |
|  | *Not confident (total)* | *27* | ***81.8*** |
|  | *Confident (total)* | *6* | *18.2* |
|  | *Total* | *33* | *100* |
| Should they be combined with other devices assessing complementary data (e.g., heart rate, O2, cardiopulmonary coupling (CPC), etc. | Yes | 28 | **80** |
|  | No | 7 | 20 |
|  | *Total* | *35* | *100* |
| **EEG-based devices** | | | |
| How confident are you in the validity of new EEG-based wearables to replace full PSGs? | Not confident at all | 3 | 10.3 |
|  | Not confident | 13 | 44.8 |
|  | Confident | 12 | 41.4 |
|  | Very confident | 1 | 3.5 |
|  | *Not confident (total)* | *16* | *55.1* |
|  | *Confident (total)* | *13* | *44.9* |
|  | *Total* | *29* | *100* |
| How confident are you in using oximeters for use at home to assess obstructive sleep apnea in cognitively unimpaired older adults? | Not confident | 5 | 16.7 |
|  | Confident | 20 | 66.7 |
|  | Very confident | 5 | 16.7 |
|  | *Confident (total)* | *25* | ***83.3*** |
|  | *Total* | *30* | *100* |
| How confident are you in the validity of such devices in cognitively unimpaired older adults? | Not confident | 6 | 20.7 |
|  | Confident | 17 | 58.6 |
|  | Very confident | 6 | 20.7 |
|  | *Confident (total)* | *23* | ***79.3*** |
|  | *Total* | *29* | *100* |
| How confident are you in the validity of such devices in participants with MCI/dementia? | Not confident at all | 1 | 3.5 |
|  | Not confident | 9 | 31 |
|  | Confident | 17 | 58.6 |
|  | Very confident | 2 | 6.9 |
|  | *Not confident (total)* | *10* | *34.4* |
|  | *Confident (total)* | *19* | *65.5* |
|  | *Total* | *29* | *100* |
| Should they be combined with other devices assessing complementary aspects (e.g., heart rate, O2…) | Yes | 29 | **85.3** |
|  | No | 5 | 14.7 |
|  | *Total* | *34* | *100* |

Results of the vote are expressed in terms of frequency (n) and percentage (%). Percentages highlighted in bold reached the 70% consensus threshold. *Abbreviations: EEG, electroencephalography; MCI, Mild Cognitive Impairment; PSG, polysomnography.*

**Supplementary Table 13. Confidence in new movement-based wearables and nearables for measuring specific sleep and circadian features in a research context.**

| **Features** | **Confidence** | **Research-quality accelerometer devices** (e.g., Philips Actiwatch, ActigraphwGT3X-BT, GeneActiv, ActTrust) | | **Consumer-quality accelerometer devices** (e.g., Oura ring, Evie Ring, Apple Watch, WHOOP, Fitbit, Garmin) | |
| --- | --- | --- | --- | --- | --- |
|  |  | **n** | **%** | **n** | **%** |
| Sleep onset latency | Confident | 12 | 40.0 | 3 | 10.0 |
|  | Not confident | 18 | 60.0 | 27 | **90.0** |
| Total sleep time | Confident | 27 | **87.1** | 13 | 41.9 |
|  | Not confident | 4 | 12.9 | 18 | 58.1 |
| Sleep onset and offset | Confident | 22 | **71.0** | 9 | 29.0 |
|  | Not confident | 9 | 29.0 | 22 | **71.0** |
| Sleep efficiency | Confident | 25 | **83.3** | 8 | 26.7 |
|  | Not confident | 5 | 16.7 | 22 | **73.3** |
| Wake after sleep onset | Confident | 26 | **83.9** | 12 | 40.0 |
|  | Not confident | 5 | 16.1 | 18 | 60.0 |
| Daytime activity, including napping | Confident | 21 | **70.0** | 8 | 26.7 |
|  | Not confident | 9 | 30.0 | 22 | **73.3** |
| Circadian features | Confident | 27 | **90.0** | 11 | 35.5 |
|  | Not confident | 3 | 10.0 | 20 | 64.5 |

Results of the vote are expressed in terms of frequency (n) and percentage (%). Percentages highlighted in bold reached the 70% consensus threshold.

**Supplementary Table 14. Confidence in new EEG-based wearables – Delphi round 2.**

| **Confidence** | **Research-quality devices**  (e.g., Dreem, Sleep Profiler,SleepImage, Accurable, Philips Actiwatch, wGT3X-BT actigraph) | | **Consumer-quality devices**  (e.g., Fitbit, Apple Watch, Oura ring, Withings Sleep Tracking Mat, WHOOP, Muse S headband, Garmin Vivosmart 4). | |
| --- | --- | --- | --- | --- |
|  | **n** | **%** | **n** | **%** |
| **How confident are you in the validity of wearable and nearable devices in MCI/AD dementia?** | | | | |
| Confident | 22 | **71.0** | 4 | 13.3 |
| Not Confident | 9 | 29.0 | 26 | **86.7** |
| **How confident are you in the validity of new EEG-based wearables to replace full PSGs for research purposes?** | | | | |
| Confident | 13 | 43.3 | 5 | 16.7 |
| Not Confident | 17 | 56.7 | 25 | **83.3** |

Results of the vote are expressed in terms of frequency (n) and percentage (%). Percentages highlighted in bold reached the 70% consensus threshold. *Abbreviations: AD, Alzheimer’s disease; EEG, electroencephalography; MCI, Mild Cognitive Impairment; PSG, polysomnography.*

**Supplementary Table 15. Identification of understudied sleep and circadian features – Delphi round 1.**

| **Feature** | | **Not important** | **Minimally important** | **Moderately important** | **Strongly important** | ***Not important (total)*** | ***Important (total)*** | ***Total*** |
| --- | --- | --- | --- | --- | --- | --- | --- | --- |
| Global self-reported measures (e.g., PSQI) | n | 6 | 14 | 10 | 4 | *20* | *14* | *34* |
|  | % | 17.6 | 41.2 | 29.4 | 11.8 | *58.8* | *41.2* | *100* |
| Excessive daytime sleepiness (e.g., ESS) | n | 4 | 8 | 18 | 6 | *12* | *24* | *36* |
|  | % | 11.1 | 22.2 | 50 | 16.7 | *33.3* | *66.7* | *100* |
| Obstructive sleep apnea | n | 0 | 2 | 6 | 26 | *2* | *32* | *34* |
|  | % | 0 | 5.9 | 17.6 | 76.5 | *5.9* | ***94.1*** | *100* |
| Parasomnias/RBD | n | 0 | 9 | 10 | 13 | *9* | *23* | *32* |
|  | % | 0 | 28.1 | 31.3 | 40.6 | *28.1* | ***71.9*** | *100* |
| Sleep-related movement disorders (e.g., PLMD) | n | 0 | 11 | 14 | 7 | *11* | *21* | *32* |
|  | % | 0 | 34.4 | 43.8 | 21.9 | *34.4* | *65.7* | *100* |
| Hypersomnolence disorders | n | 0 | 13 | 11 | 4 | *13* | *15* | *28* |
|  | % | 0 | 46.4 | 39.3 | 14.3 | *46.4* | *53.6* | *100* |
| Insomnia | n | 0 | 3 | 10 | 21 | *3* | *31* | *34* |
|  | % | 0 | 8.8 | 29.4 | 61.8 | *8.8* | ***91.2*** | *100* |
| General actigraphy features | n | 0 | 2 | 25 | 7 | *2* | *32* | *34* |
|  | % | 0 | 5.9 | 73.5 | 20.6 | *5.9* | ***94.1*** | *100* |
| Actigraphy-defined sleep variability | n | 0 | 2 | 19 | 13 | *2* | *32* | *34* |
|  | % | 0 | 5.9 | 55.9 | 38.2 | *5.9* | ***94.1*** | *100* |
| Harmonization of actigraphy methodologies | n | 0 | 0 | 12 | 22 | *0* | *34* | *34* |
|  | % | 0 | 0 | 35.3 | 64.7 | *0* | ***100*** | *100* |
| Gold-standard circadian outputs | n | 0 | 1 | 10 | 22 | *1* | *32* | *33* |
|  | % | 0 | 3 | 30.3 | 66.7 | *3* | ***97*** | *100* |
| Naps | n | 0 | 6 | 13 | 15 | *6* | *28* | *34* |
|  | % | 0 | 17.6 | 38.2 | 44.1 | *17.6* | ***82.3*** | *100* |
| Sleep cycles | n | 0 | 5 | 19 | 9 | *5* | *28* | *33* |
|  | % | 0 | 15.2 | 57.6 | 27.3 | *15.2* | ***84.9*** | *100* |
| Sleep duration | n | 0 | 5 | 19 | 12 | *5* | *31* | *36* |
|  | % | 0 | 13.9 | 52.8 | 33.3 | *13.9* | ***86.1*** | *100* |
| Sleep fragmentation | n | 0 | 2 | 14 | 20 | *2* | *34* | *36* |
|  | % | 0 | 5.6 | 38.9 | 55.6 | *5.6* | ***94.5*** | *100* |
| N1 sleep | n | 0 | 11 | 14 | 5 | *11* | *19* | *30* |
|  | % | 0 | 36.7 | 46.7 | 16.7 | *36.7* | *63.4* | *100* |
| N2 sleep | n | 0 | 5 | 15 | 10 | *5* | *25* | *30* |
|  | % | 0 | 16.7 | 50 | 33.3 | *16.7* | ***83.3*** | *100* |
| N3 sleep / slow wave sleep | n | 0 | 1 | 10 | 22 | *1* | *32* | *33* |
|  | % | 0 | 3 | 30.3 | 66.7 | *3* | ***97*** | *100* |
| REM sleep | n | 0 | 1 | 13 | 20 | *1* | *33* | *34* |
|  | % | 0 | 2.9 | 38.2 | 58.8 | *2.9* | ***97.1*** | *100* |
| Spectral analyses/qEEG | n | 0 | 1 | 11 | 19 | *1* | *30* | *31* |
|  | % | 0 | 3.2 | 35.5 | 61.3 | *3.2* | ***96.8*** | *100* |
| EEG connectivity/coupling | n | 0 | 1 | 14 | 15 | *1* | *29* | *30* |
|  | % | 0 | 3.3 | 46.7 | 50 | *3.3* | ***96.7*** | *100* |
| Sleep spindles | n | 0 | 0 | 12 | 19 | *0* | *31* | *31* |
|  | % | 0 | 0 | 38.7 | 61.3 | *0* | ***100*** | *100* |
| Slow waves | n | 0 | 0 | 9 | 25 | *0* | *34* | *34* |
|  | % | 0 | 0 | 26.5 | 73.5 | *0* | ***100*** | *100* |
| K complexes | n | 1 | 5 | 11 | 11 | *6* | *22* | *28* |
|  | % | 3.6 | 17.9 | 39.3 | 39.3 | *21.5* | ***78.6*** | *100* |
| Epilepsy spikes characteristics | n | 1 | 5 | 12 | 8 | *6* | *20* | *26* |
|  | % | 3.8 | 19.2 | 46.2 | 30.8 | *23* | ***77*** | *100* |

Results of the vote are expressed in terms of frequency (n) and percentage (%). Percentages highlighted in bold reached the 70% consensus threshold. *Abbreviations: EEG, electroencephalography; ESS, Epworth Sleepiness Scale; PLMD, periodic limb movement disorder; PSQI, Pittsburgh Sleep Quality Index; qEEG, quantitative EEG; REM, rapid eye movement; RBD, REM sleep behavior disorder.*

**Supplementary Table 16. Identification of understudied populations – Delphi round 1.**

| **Answer options** | | **Not important** | **Minimally important** | **Moderately important** | **Strongly important** | ***Not important (total)*** | ***Important (total)*** | ***Total*** |
| --- | --- | --- | --- | --- | --- | --- | --- | --- |
| Stratification according to biological sex | n | 0 | 5 | 10 | 20 | *5* | *30* | *35* |
|  | % | 0 | 14.3 | 28.6 | 57.1 | *14.3* | ***85.7*** | *100* |
| Individuals with genetic risk factors for AD (e.g., APOE ε4 carriers) | n | 0 | 2 | 10 | 24 | *2* | *34* | *36* |
|  | % | 0 | 5.6 | 27.8 | 66.7 | *5.6* | ***94.5*** | *100* |
| Extreme chronotypes | n | 0 | 4 | 20 | 10 | *4* | *30* | *34* |
|  | % | 0 | 11.8 | 58.8 | 29.4 | *11.8* | ***88.2*** | *100* |
| Short sleepers (<6h) | n | 0 | 4 | 20 | 13 | *4* | *33* | *37* |
|  | % | 0 | 10.8 | 54.1 | 35.1 | *10.8* | ***89.2*** | *100* |
| Long sleepers (>9h) | n | 0 | 4 | 22 | 10 | *4* | *32* | *36* |
|  | % | 0 | 11.1 | 61.1 | 27.8 | *11.1* | ***88.9*** | *100* |
| People with sleep disorders | n | 0 | 1 | 11 | 24 | *1* | *35* | *36* |
|  | % | 0 | 2.8 | 30.6 | 66.7 | *2.8* | ***97.3*** | *100* |
| Individuals with shift work history | n | 0 | 2 | 14 | 19 | *2* | *33* | *35* |
|  | % | 0 | 5.7 | 40 | 54.3 | *5.7* | ***94.3*** | *100* |
| Individuals with mental health disorders | n | 0 | 2 | 19 | 14 | *2* | *33* | *35* |
|  | % | 0 | 5.7 | 54.3 | 40 | *5.7* | ***94.3*** | *100* |
| Ethnical disparities | n | 1 | 4 | 14 | 17 | *5* | *31* | *36* |
|  | % | 2.8 | 11.1 | 38.9 | 47.2 | *13.9* | ***86.1*** | *100* |
| Non-AD dementias | n | 0 | 1 | 12 | 23 | *1* | *35* | *36* |
|  | % | 0 | 2.8 | 33.3 | 63.9 | *2.8* | ***97.2*** | *100* |
| People in low- and middle-income countries | n | 0 | 4 | 17 | 14 | *4* | *31* | *35* |
|  | % | 0 | 11.4 | 48.6 | 40 | *11.4* | ***88.6*** | *100* |
| Middle aged groups (40-65 years old) | n | 0 | 2 | 13 | 21 | *2* | *34* | *36* |
|  | % | 0 | 5.6 | 36.1 | 58.3 | *5.6* | ***94.4*** | *100* |
| Older groups (>65 years old) | n | 0 | 3 | 14 | 20 | *3* | *34* | *37* |
|  | % | 0 | 8.1 | 37.8 | 54.1 | *8.1* | ***91.9*** | *100* |
| Other comments: patients on benzodiazepines (n=1), perimenopausal period (n=1) | | | | | | | | |

Results of the vote are expressed in terms of frequency (n) and percentage (%). The ‘Not Important’ total was computed by summing the frequencies of the 'Not Important' and 'Minimally Important’ options, while the ‘Important’ total represents the sum of frequencies of the 'Moderately Important' and 'Strongly Important' options. Percentages highlighted in bold reached the 70% consensus threshold. *Abbreviations: AD, Alzheimer’s disease; APOE, apolipoprotein E; h, hour.*
