## Appendix A for "International recommendations for sleep and circadian research in aging and Alzheimer’s disease: a Delphi consensus study"

CONSENSUS RECOMMENDATIONS ON RELEVANT SLEEP AND CIRCADIAN RHYTHMS MEASURES IN THE CONTEXT OF AGING AND ALZHEIMER’S DISEASE – **DELPHI SURVEY (ROUND 1).**

1. **Section 1: General information**
   1.1 Name
   1.2 Email address
   1.3 Primary affiliation(s)/Institution
   1.4 Country
   1.5 Postcode
   1.6 Professional setting *(Please tick all that apply)*

- **Clinician**
  - Neurologist
  - Geriatrician
  - Psychiatrist
  - Pulmonary physicians
  - Otolaryngologists
  - Anesthesiologists
  - General Practitioner (GP)/Primary Care Provider (PCP)
  - Neuropsychologist
  - Nurse practitioner (NP)
  - Physician assistant (PA)
  - Clinical psychologist
  - Occupational therapist
  - Sleep physician
  - Sleep technician
  - Other, *please specify*

1.6.1	Are you still in clinical practice? Yes/No

1.6.2	Years in clinical practice, *please specify:*

- **Researcher**
  - Sleep medicine
  - Sleep neurophysiology
  - Alzheimer’s and dementia biomarkers,
  - …choose

Fluid biomarkers

Genetics

Imaging

- Epidemiology/Public health
- Health services
- Basic science
- Clinical or applied research
- Clinical trials
- Engineering or computer science
- Data science/AI
- Industry: Sleep technology company
- Industry: Pharmaceutical company
- Industry: Other
- Other research field, *please specify*

1.6.3	Are you still conducting research? Yes/No

1.6.4	Years of research experience, *please specify: (open field)*

*1.6.5	Are you an early career researcher (defined as <8 years since PhD)? Yes/No.*

1.7 Do you have any direct or indirect conflicts of interest relevant to the scope of this work (include consultancies, honoraria, company shares, or employment in industry)?

Yes/No

If Yes, please describe all relevant conflicts of interest hereafter

| Consultancies | *Open field* |
| --- | --- |
| Honoraria | *Open field* |
| Company shares | *Open field* |
| Employment in industry | *Open field* |
| Other, please specify | *Open field* |

1.8 Highest qualification
- Masters degree
- Medical degree (MD)
- PhD or equivalent
- Medical degree and PhD (e.g., MD, PhD)
- Medical degree and Masters degree (e.g., MD, MSc)
- Certified sleep technologist
- Other, *please specify*
1.9 Current primary position
    - PhD student/candidate
    - Postdoctoral Fellow
    - Faculty Academic appointment (e.g., Dr, Asst/Prof, Assoc.Prof, Prof) or full-time/tenured researcher
    - Clinical/medical hospital appointment (medical staff)
    - Director of institute
    - Company director
    - Employee of company/industry
    - Patient and public sleep advocacy group
    - Retired *(please specify your past primary position in the open field below)*
    - Other, *please specify*
1.10 Working environment
     - Hospital or health service (outpatient facility)
     - Primary care
     - Private practice
     - Medical school
     - University
     - Research center or institute
     - Industry
     - Other, *please specify*
1.11 Are you from a low- or middle-income country? *Yes/No*
1.12 What is your race/ethnicity?
     - American Indian or Alaskan Native
     - Asian
     - Black/African American
     - Hispanic/Latino
     - Native Hawaiian or Pacific Islander
     - White
     - Two or more races or ethnicities
     - Prefer not to answer
     - Prefer to self-describe

*Please self-describe:*

1.13 What is your primary language?
1.14 What is your secondary language?

2. **Section 2: Relevant sleep and circadian features**

***Answer options:***

***1: Strongly disagree; 2: disagree; 3: agree; 4: strongly agree, N/A: unable to answer.***

2.1. **Relevant features to characterize pathological ageing**
     2.1.1. **In your experience, which sleep and circadian features are sensitive to preclinical AD, MCI and AD dementia compared to normal aging?**

*NB: “Preclinical AD” here refers to cognitively unimpaired individuals who are engaged in the Alzheimer’s continuum (i.e., amyloid-positive at a minimum).*

| *Variable* | *Preclinical AD* | *MCI* | *AD dementia* |
| --- | --- | --- | --- |
| **Clinical symptoms and self-reported data** | | | |
| General sleep disturbance/poor sleep quality | **1 2 3 4 N/A** | **1 2 3 4 N/A** | **1 2 3 4 N/A** |
| Excessive daytime sleepiness | **1 2 3 4 N/A** | **1 2 3 4 N/A** | **1 2 3 4 N/A** |
| Sleep-related breathing disorders (e.g., OSA) symptoms | **1 2 3 4 N/A** | **1 2 3 4 N/A** | **1 2 3 4 N/A** |
| Parasomnias (e.g., RBD) and related symptoms | **1 2 3 4 N/A** | **1 2 3 4 N/A** | **1 2 3 4 N/A** |
| Sleep-related movement disorders (e.g., PLMD) | **1 2 3 4 N/A** | **1 2 3 4 N/A** | **1 2 3 4 N/A** |
| Hypersomnolence disorders (e.g., narcolepsy, hypersomnia) | **1 2 3 4 N/A** | **1 2 3 4 N/A** | **1 2 3 4 N/A** |
| Insomnia | **1 2 3 4 N/A** | **1 2 3 4 N/A** | **1 2 3 4 N/A** |
| Sleep timing (i.e., advanced or delayed sleep phase) | **1 2 3 4 N/A** | **1 2 3 4 N/A** | **1 2 3 4 N/A** |
| Self-reported sleep onset latency (SOL) | **1 2 3 4 N/A** | **1 2 3 4 N/A** | **1 2 3 4 N/A** |
| Self-reported total sleep time (TST) | **1 2 3 4 N/A** | **1 2 3 4 N/A** | **1 2 3 4 N/A** |
| Self-reported sleep efficiency (SE) | **1 2 3 4 N/A** | **1 2 3 4 N/A** | **1 2 3 4 N/A** |
| Self-reported nocturnal awakenings (e.g., number or duration) | **1 2 3 4 N/A** | **1 2 3 4 N/A** | **1 2 3 4 N/A** |
| Self-reported napping | **1 2 3 4 N/A** | **1 2 3 4 N/A** | **1 2 3 4 N/A** |
| **From polysomnography** | | | |
| *Sleep macroarchitecture features* |  |  |  |
| Sleep onset latency (SOL) | **1 2 3 4 N/A** | **1 2 3 4 N/A** | **1 2 3 4 N/A** |
| Total sleep time (TST) | **1 2 3 4 N/A** | **1 2 3 4 N/A** | **1 2 3 4 N/A** |
| Sleep efficiency (SE) | **1 2 3 4 N/A** | **1 2 3 4 N/A** | **1 2 3 4 N/A** |
| Sleep fragmentation (e.g., number of awakenings, arousals, stages shifts) | **1 2 3 4 N/A** | **1 2 3 4 N/A** | **1 2 3 4 N/A** |
| Wake after sleep onset (WASO) | **1 2 3 4 N/A** | **1 2 3 4 N/A** | **1 2 3 4 N/A** |
| REM sleep latency | **1 2 3 4 N/A** | **1 2 3 4 N/A** | **1 2 3 4 N/A** |
| Duration in each sleep stage/% | **1 2 3 4 N/A** | **1 2 3 4 N/A** | **1 2 3 4 N/A** |
| *Breathing features* |  |  |  |
| Apnea Hypopnea Index (AHI) | **1 2 3 4 N/A** | **1 2 3 4 N/A** | **1 2 3 4 N/A** |
| Oxygen Desaturation Index (ODI) | **1 2 3 4 N/A** | **1 2 3 4 N/A** | **1 2 3 4 N/A** |
| *Sleep microarchitecture features* |  |  |  |
| NREM sleep spectral power (including slow wave activity) | **1 2 3 4 N/A** | **1 2 3 4 N/A** | **1 2 3 4 N/A** |
| REM sleep spectral power (including REM sleep EEG slowing) | **1 2 3 4 N/A** | **1 2 3 4 N/A** | **1 2 3 4 N/A** |
| Sleep spindles characteristics | **1 2 3 4 N/A** | **1 2 3 4 N/A** | **1 2 3 4 N/A** |
| Slow waves characteristics | **1 2 3 4 N/A** | **1 2 3 4 N/A** | **1 2 3 4 N/A** |
| K complexes | **1 2 3 4 N/A** | **1 2 3 4 N/A** | **1 2 3 4 N/A** |
| Epileptiform EEG activity | **1 2 3 4 N/A** | **1 2 3 4 N/A** | **1 2 3 4 N/A** |
| EEG coupling/connectivity meaures | **1 2 3 4 N/A** | **1 2 3 4 N/A** | **1 2 3 4 N/A** |
| *Features consistent with REM Sleep Behavior Disorder* | **1 2 3 4 N/A** | **1 2 3 4 N/A** | **1 2 3 4 N/A** |
| **From actigraphy (± associated diary)** | | | |
| Sleep onset latency (SOL) | **1 2 3 4 N/A** | **1 2 3 4 N/A** | **1 2 3 4 N/A** |
| Total sleep time (TST) | **1 2 3 4 N/A** | **1 2 3 4 N/A** | **1 2 3 4 N/A** |
| Sleep onset and offset | **1 2 3 4 N/A** | **1 2 3 4 N/A** | **1 2 3 4 N/A** |
| Sleep efficiency | **1 2 3 4 N/A** | **1 2 3 4 N/A** | **1 2 3 4 N/A** |
| Wake after sleep onset (WASO) | **1 2 3 4 N/A** | **1 2 3 4 N/A** | **1 2 3 4 N/A** |
| Daytime activity, including potential napping | **1 2 3 4 N/A** | **1 2 3 4 N/A** | **1 2 3 4 N/A** |
| Other features derived from non-parametric or cosinor analysis | **1 2 3 4 N/A** | **1 2 3 4 N/A** | **1 2 3 4 N/A** |
| **Other, please specify** *(open field)* | **1 2 3 4 N/A** | **1 2 3 4 N/A** | **1 2 3 4 N/A** |

2.1.2. **Do you use actigraphy and PSG recordings in your research setting?** *Please tick all that apply.*
- Actigraphy and other “research-quality” accelerometer wearable devices (e.g., Actigraph, GENEActiv, etc.)
- Commercially-available accelerometer wearable devices (e.g., Fitbit, Apple Watch, etc.)
- PSG/EEG-based wearables
- Oximetry monitors (e.g., WatchPat, WristOx…)
- None
- Other, please specify *(open field)*
  2.1.3. **How many days of actigraphy recordings are ideal in a research setting?**
- <3 days
- 3 days
- 5 days
- 7 days
- 10 days
- 14 days
- >14 days
- Unable to answer

3. **Section 3: Recommendations on data acquisition and report**
   3.1. **Which method do you feel is best to assess the following features in each clinical group?**

| Measure | Preclinical AD | MCI | AD dementia |
| --- | --- | --- | --- |
| Time in Bed (TIB) | - Patient self-report - Informant report - Accelerometer / wearable devices - In-lab PSG/EEG - At-home PSG/EEG - Other, please specify | - Patient self-report - Informant report - Accelerometer / wearable devices - In-lab PSG/EEG - At-home PSG/EEG - Other, please specify | - Patient self-report - Informant report - Accelerometer / wearable devices - In-lab PSG/EEG - At-home PSG/EEG - Other, please specify |
| Sleep duration/total sleep time (TST) | - Patient self-report - Informant report - Accelerometer /wearable devices - In-lab PSG/EEG - At-home PSG/EEG - Other, please specify | - Patient self-report - Informant report - Accelerometer /wearable devices - In-lab PSG/EEG - At-home PSG/EEG - Other, please specify | - Patient self-report - Informant report - Accelerometer /wearable devices - In-lab PSG/EEG - At-home PSG/EEG - Other, please specify |
| Sleep onset latency (SOL) | - Patient self-report - Informant report - Accelerometer wearable devices - In-lab PSG/EEG - At-home PSG/EEG - Other, please specify | - Patient self-report - Informant report - Accelerometer wearable devices - In-lab PSG/EEG - At-home PSG/EEG - Other, please specify | - Patient self-report - Informant report - Accelerometer wearable devices - In-lab PSG/EEG - At-home PSG/EEG - Other, please specify |
| Sleep efficiency | - Patient self-report - Informant report - Accelerometer wearable devices - In-lab PSG/EEG - At-home PSG/EEG - Other, please specify | - Patient self-report - Informant report - Accelerometer wearable devices - In-lab PSG/EEG - At-home PSG/EEG - Other, please specify | - Patient self-report - Informant report - Accelerometer wearable devices - In-lab PSG/EEG - At-home PSG/EEG - Other, please specify |
| Sleep fragmentation (e.g., awakenings) | - Patient self-report - Informant report - Accelerometer wearable devices - In-lab PSG/EEG - At-home PSG/EEG - Other, please specify | - Patient self-report - Informant report - Accelerometer wearable devices - In-lab PSG/EEG - At-home PSG/EEG - Other, please specify | - Patient self-report - Informant report - Accelerometer wearable devices - In-lab PSG/EEG - At-home PSG/EEG - Other, please specify |
| Wake after sleep onset (WASO) | - Patient self-report - Informant report - Accelerometer wearable devices - In-lab PSG/EEG - At-home PSG/EEG - Other, please specify | - Patient self-report - Informant report - Accelerometer wearable devices - In-lab PSG/EEG - At-home PSG/EEG - Other, please specify | - Patient self-report - Informant report - Accelerometer wearable devices - In-lab PSG/EEG - At-home PSG/EEG - Other, please specify |
| Naps (timing, duration, frequency) | - Patient self-report - Informant report - Accelerometer wearable devices - In-lab PSG/EEG - At-home PSG/EEG - Other, please specify | - Patient self-report - Informant report - Accelerometer wearable devices - In-lab PSG/EEG - At-home PSG/EEG - Other, please specify | - Patient self-report - Informant report - Accelerometer wearable devices - In-lab PSG/EEG - At-home PSG/EEG - Other, please specify |
| Sleep-disordered breathing | - Patient self-report - Informant report - At home oximetry monitors - In-lab PSG/sleep apnea test - At-home PSG/ sleep apnea test - Other, please specify | - Patient self-report - Informant report - At home oximetry monitors - In-lab PSG/sleep apnea test - At-home PSG/ sleep apnea test - Other, please specify | - Patient self-report - Informant report - At home oximetry monitors - In-lab PSG/sleep apnea test - At-home PSG/ sleep apnea test - Other, please specify |

3.2. **Consensus on the minimum standards for data collection and analysis in future studies/clinical trials**
     3.2.1. **Recruitment and characterization of participants:**

In addition to age and biological sex, which characteristics are the most essential to collect and report in sleep studies in the context of aging and dementia?

| **Demographics, clinical and biological features** | |
| --- | --- |
| Gender | - Absolutely essential (the findings are inconclusive or biased if this factor is not controlled for or reported) - Necessary for a well-defined study - Not essential/optional - Unable to answer |
| Cognitive diagnosis | - Absolutely essential (the findings are inconclusive or biased if this factor is not controlled for or reported). - Necessary for a well-defined study. - Not essential/optional. - Unable to answer |
| Subjective cognitive/memory concerns | - Absolutely essential (the findings are inconclusive or biased if this factor is not controlled for or reported) - Necessary for a well-defined study - Not essential/optional - Unable to answer |
| Biomarker profile for clinical staging or ATN classification | - Absolutely essential (the findings are inconclusive or biased if this factor is not controlled for or reported) - Necessary for a well-defined study - Not essential/optional. - Unable to answer |
| Race | - Absolutely essential (the findings are inconclusive or biased if this factor is not controlled for or reported) - Necessary for a well-defined study - Not essential/optional - Unable to answer |
| Ethnicity | - Absolutely essential (the findings are inconclusive or biased if this factor is not controlled for or reported) - Necessary for a well-defined study - Not essential/optional - Unable to answer |
| Education level | - Absolutely essential (the findings are inconclusive or biased if this factor is not controlled for or reported) - Necessary for a well-defined study - Not essential/optional - Unable to answer |
| Living arrangement/ Residential setting | - Absolutely essential (the findings are inconclusive or biased if this factor is not controlled for or reported) - Necessary for a well-defined study - Not essential/optional - Unable to answer |
| Social network | - Absolutely essential (the findings are inconclusive or biased if this factor is not controlled for or reported) - Necessary for a well-defined study - Not essential/optional - Unable to answer |
| Employment | - Absolutely essential (the findings are inconclusive or biased if this factor is not controlled for or reported) - Necessary for a well-defined study - Not essential/optional |
| Vocational status (e.g., retirement) | - Absolutely essential (the findings are inconclusive or biased if this factor is not controlled for or reported) - Necessary for a well-defined study - Not essential/optional - Unable to answer |
| Caring responsibilities | - Absolutely essential (the findings are inconclusive or biased if this factor is not controlled for or reported) - Necessary for a well-defined study - Not essential/optional - Unable to answer |
| APOE ε4 | - Absolutely essential (the findings are inconclusive or biased if this factor is not controlled for or reported) - Necessary for a well-defined study - Not essential/optional - Unable to answer |
| Body mass index | - Absolutely essential (the findings are inconclusive or biased if this factor is not controlled for or reported) - Necessary for a well-defined study - Not essential/optional - Unable to answer |
| Blood pressure | - Absolutely essential (the findings are inconclusive or biased if this factor is not controlled for or reported) - Necessary for a well-defined study - Not essential/optional - Unable to answer |
| Lifestyle: physical activity levels | - Absolutely essential (the findings are inconclusive or biased if this factor is not controlled for or reported) - Necessary for a well-defined study - Not essential/optional - Unable to answer |
| Lifestyle: diet | - Absolutely essential (the findings are inconclusive or biased if this factor is not controlled for or reported) - Necessary for a well-defined study - Not essential/optional - Unable to answer |
| Coffee, tea, energetic drinks consumption | - Absolutely essential (the findings are inconclusive or biased if this factor is not controlled for or reported) - Necessary for a well-defined study - Not essential/optional - Unable to answer |
| Daily light exposure | - Absolutely essential (the findings are inconclusive or biased if this factor is not controlled for or reported) - Necessary for a well-defined study - Not essential/optional - Unable to answer |
| Season (of data collection/clinical assessment) | - Absolutely essential (the findings are inconclusive or biased if this factor is not controlled for or reported) - Necessary for a well-defined study - Not essential/optional - Unable to answer |
| Sleeping arrangement | - Absolutely essential (the findings are inconclusive or biased if this factor is not controlled for or reported) - Necessary for a well-defined study - Not essential/optional - Unable to answer |
| Number of pregnancies | - Absolutely essential (the findings are inconclusive or biased if this factor is not controlled for or reported) - Necessary for a well-defined study - Not essential/optional - Unable to answer |
| Menopause: current status | - Absolutely essential (the findings are inconclusive or biased if this factor is not controlled for or reported) - Necessary for a well-defined study - Not essential/optional - Unable to answer |
| Menopause: age of onset | - Absolutely essential (the findings are inconclusive or biased if this factor is not controlled for or reported) - Necessary for a well-defined study - Not essential/optional - Unable to answer |
| **Comorbidities (current and/or history)** | |
| Smoking | - Absolutely essential (the findings are inconclusive or biased if this factor is not controlled for or reported) - Necessary for a well-defined study - Not essential/optional - Unable to answer |
| Alcohol intake | - Absolutely essential (the findings are inconclusive or biased if this factor is not controlled for or reported) - Necessary for a well-defined study - Not essential/optional - Unable to answer |
| Cannabis/THC | - Absolutely essential (the findings are inconclusive or biased if this factor is not controlled for or reported) - Necessary for a well-defined study - Not essential/optional - Unable to answer |
| Illicit substances | - Absolutely essential (the findings are inconclusive or biased if this factor is not controlled for or reported) - Necessary for a well-defined study - Not essential/optional - Unable to answer |
| State anxiety | - Absolutely essential (the findings are inconclusive or biased if this factor is not controlled for or reported) - Necessary for a well-defined study - Not essential/optional - Unable to answer |
| Trait anxiety | - Absolutely essential (the findings are inconclusive or biased if this factor is not controlled for or reported) - Necessary for a well-defined study - Not essential/optional - Unable to answer |
| Depressive symptoms | - Absolutely essential (the findings are inconclusive or biased if this factor is not controlled for or reported) - Necessary for a well-defined study - Not essential/optional - Unable to answer |
| Cardiovascular diseases | - Absolutely essential (the findings are inconclusive or biased if this factor is not controlled for or reported) - Necessary for a well-defined study - Not essential/optional - Unable to answer |
| Neurological diseases (e.g., stroke, epilepsy, Parkinsons) | - Absolutely essential (the findings are inconclusive or biased if this factor is not controlled for or reported) - Necessary for a well-defined study - Not essential/optional - Unable to answer |
| Psychiatric disorders and history of mental illness | - Absolutely essential (the findings are inconclusive or biased if this factor is not controlled for or reported) - Necessary for a well-defined study - Not essential/optional - Unable to answer |
| Metabolic syndromes/disorders | - Absolutely essential (the findings are inconclusive or biased if this factor is not controlled for or reported) - Necessary for a well-defined study - Not essential/optional - Unable to answer |
| Cancer | - Absolutely essential (the findings are inconclusive or biased if this factor is not controlled for or reported) - Necessary for a well-defined study - Not essential/optional - Unable to answer |
| Traumatic Brain Injury | - Absolutely essential (the findings are inconclusive or biased if this factor is not controlled for or reported) - Necessary for a well-defined study - Not essential/optional - Unable to answer |
| **Sleep** | |
| Chronotype | - Absolutely essential (the findings are inconclusive or biased if this factor is not controlled for or reported) - Necessary for a well-defined study - Not essential/optional - Unable to answer |
| History of shift work | - Absolutely essential (the findings are inconclusive or biased if this factor is not controlled for or reported) - Necessary for a well-defined study - Not essential/optional - Unable to answer |
| Current shift work | - Absolutely essential (the findings are inconclusive or biased if this factor is not controlled for or reported) - Necessary for a well-defined study - Not essential/optional - Unable to answer |
| OSA diagnosis | - Absolutely essential (the findings are inconclusive or biased if this factor is not controlled for or reported) - Necessary for a well-defined study - Not essential/optional - Unable to answer |
| OSA treatment (e.g., CPAP use) | - Absolutely essential (the findings are inconclusive or biased if this factor is not controlled for or reported) - Necessary for a well-defined study - Not essential/optional - Unable to answer |
| Insomnia symptoms | - Absolutely essential (the findings are inconclusive or biased if this factor is not controlled for or reported) - Necessary for a well-defined study - Not essential/optional - Unable to answer |
| Insomnia treatment (e.g., CBT-I) | - Absolutely essential (the findings are inconclusive or biased if this factor is not controlled for or reported) - Necessary for a well-defined study - Not essential/optional - Unable to answer |
| Restless legs syndrome | - Absolutely essential (the findings are inconclusive or biased if this factor is not controlled for or reported) - Necessary for a well-defined study - Not essential/optional - Unable to answer |
| Other co-morbid sleep disorders *(please specify in the open field below)* | - Absolutely essential (the findings are inconclusive or biased if this factor is not controlled for or reported) - Necessary for a well-defined study - Not essential/optional - Unable to answer |
| Heart rate variability | - Absolutely essential (the findings are inconclusive or biased if this factor is not controlled for or reported) - Necessary for a well-defined study - Not essential/optional - Unable to answer |
| **Medication** | |
| Benzodiazepines | - Absolutely essential (the findings are inconclusive or biased if this factor is not controlled for or reported) - Necessary for a well-defined study - Not essential/optional - Unable to answer |
| Other benzodiazepine receptor agonist hypnotics | - Absolutely essential (the findings are inconclusive or biased if this factor is not controlled for or reported) - Necessary for a well-defined study - Not essential/optional - Unable to answer |
| Trazadone | - Absolutely essential (the findings are inconclusive or biased if this factor is not controlled for or reported) - Necessary for a well-defined study - Not essential/optional - Unable to answer |
| Other approved hypnotics (e.g., DORAs, ramelteon…) | - Absolutely essential (the findings are inconclusive or biased if this factor is not controlled for or reported) - Necessary for a well-defined study - Not essential/optional - Unable to answer |
| Antidepressants. | - Absolutely essential (the findings are inconclusive or biased if this factor is not controlled for or reported) - Necessary for a well-defined study - Not essential/optional   Unable to answer |
| Antipsychotics | - Absolutely essential (the findings are inconclusive or biased if this factor is not controlled for or reported) - Necessary for a well-defined study - Not essential/optional - Unable to answer |
| Opioids | - Absolutely essential (the findings are inconclusive or biased if this factor is not controlled for or reported) - Necessary for a well-defined study - Not essential/optional - Unable to answer |
| Antihistamines | - Absolutely essential (the findings are inconclusive or biased if this factor is not controlled for or reported) - Necessary for a well-defined study - Not essential/optional - Unable to answer |
| Acetylcholinesterase (ACE) inhibitors | - Absolutely essential (the findings are inconclusive or biased if this factor is not controlled for or reported) - Necessary for a well-defined study - Not essential/optional - Unable to answer |
| Memantine | - Absolutely essential (the findings are inconclusive or biased if this factor is not controlled for or reported) - Necessary for a well-defined study - Not essential/optional - Unable to answer |
| Monoclonal antibody | - Absolutely essential (the findings are inconclusive or biased if this factor is not controlled for or reported) - Necessary for a well-defined study - Not essential/optional - Unable to answer |
| Non-narcotic analgesics | - Absolutely essential (the findings are inconclusive or biased if this factor is not controlled for or reported) - Necessary for a well-defined study - Not essential/optional - Unable to answer |
| Narcotic analgesics | - Absolutely essential (the findings are inconclusive or biased if this factor is not controlled for or reported) - Necessary for a well-defined study - Not essential/optional - Unable to answer |
| Anticonvulsants | - Absolutely essential (the findings are inconclusive or biased if this factor is not controlled for or reported) - Necessary for a well-defined study - Not essential/optional - Unable to answer |
| Stimulants (e.g., Ritalin) | - Absolutely essential (the findings are inconclusive or biased if this factor is not controlled for or reported) - Necessary for a well-defined study - Not essential/optional - Unable to answer |
| Hormone therapy | - Absolutely essential (the findings are inconclusive or biased if this factor is not controlled for or reported) - Necessary for a well-defined study - Not essential/optional - Unable to answer |
| Melatonin | - Absolutely essential (the findings are inconclusive or biased if this factor is not controlled for or reported) - Necessary for a well-defined study - Not essential/optional - Unable to answer |
| Vitamins and supplements | - Absolutely essential (the findings are inconclusive or biased if this factor is not controlled for or reported) - Necessary for a well-defined study - Not essential/optional - Unable to answer |
| Phytotherapy and homeopathy | - Absolutely essential (the findings are inconclusive or biased if this factor is not controlled for or reported) - Necessary for a well-defined study - Not essential/optional - Unable to answer |
| **Other, please specify *(open field)*** | |

3.2.2. **Identification of current limits and areas of improvement**

***Answer options: 1: not confident at all, 2: not confident, 3: confident, 4: very confident, N/A: unable to answer (Please select this option if you are unsure or feel that an item falls out of your expertise).***

| **Questionnaires/self-report** | |
| --- | --- |
| Do you feel that the quality of self-reported data depends on the cognitive status of the patient/participant? | **1 2 3 4 (N/A)** |
| How confident are you in the reliability of single-item subjective self- or informant-reported sleep measures (e.g., “sleep disturbances” item of the NPI)? | **1 2 3 4 (N/A)** |
| How confident are you in informant-related sleep data? | **1 2 3 4 (N/A)** |
| Do you feel the reliability of informant-related sleep data depends on the cognitive status of the informant? | **1 2 3 4 (N/A)** |
| Do you feel that sleep questionnaires should be re-validated for the ageing/MCI/AD population? | **1 2 3 4 (N/A)** |
| Do you feel that the scoring of sleep questionnaires should be adapted/standardized based on the ageing /MCI or AD population? | **1 2 3 4 (N/A)** |
| Comments *(open field)* | |
| **Actigraphy/accelerometer devices** | |
| Should a sleep diary always be proposed in conjunction with actigraphy in a research context? | **1 2 3 4 (N/A)** |
| What questions should be asked in a sleep diary? | - Sleep latency - Use of caffeine - Use of alcohol - Types of sleep disruptions - Wake time - Feeling refreshed upon waking - Other, please specify |
| How confident are you in sleep diary data as a tool for quality control? How much do you rely on it to interpret your recordings? | **1 2 3 4 (N/A)** |
| Do you alter the actigraphy recording based on diary report? | - Yes - No - Sometimes - N/A |
| Would you recommend using software-implemented algorithms or in-house processing for actigraphy? | - Software-implemented - Please specify - In-house - Both - Unable to answer   Comments *(open field)* |
| **PSG/EEG-based wearables** | |
| What minimum EEG montage do you recommend in a research setting *(please tick all that apply)*? | - Fp1 - Fp2 - F3 - F4 - F7 - F8 - Fz - T3 - T4 - T5 - T6 - C3 - C4 - Cz - P3 - P4 - Pz - O1 - O2 - Other, please specify *(open field)* - Unable to answer |
| For optimal compromise between feasibility and ecological validity, would you favor in-home or in-lab settings for future protocols and clinical trials? | - Always in-lab PSG regardless of the clinical group and research question - Always at-home PSG regardless of the clinical group and research question - The choice depends on the research question only - The choice depends on the clinical group only - The choice depends on both the question and the clinical group - Unable to answer |
| For cohort studies, do you feel a habituation night should be systematically performed? | - No - Yes, always (regardless of the clinical group) - Only in cognitively unimpaired participants, but not MCI/dementia patients - Unable to answer |
| Who should be excluded or considered in sensitivity analyses in studies examining the mechanistic link between sleep and dementia? | - People with stroke - People with other comorbid medical, neurological or psychiatric diseases - People with moderate to severe OSA - People with traumatic brain injury - Depends on the study - Other, please specify *(open field)* |
| In the design of clinical trials, over what timeframe should we measure prior sleep and/or psychotropic medication use? | - 1 week - 2 weeks - 1 month - Other, please specify *(open field)* - Unable to answer |

3.3. **Confidence on the use of newly available devices.**

***Answer options: 1: not confident at all, 2: not confident, 3: confident, 4: very confident, N/A: unable to answer (Please select this option if you are unsure or feel that an item falls out of your expertise).***

| **Actigraphy/accelerometer devices** | |
| --- | --- |
| How confident are you in movement-based devices recording movement only? | **1 2 3 4 (N/A)** |
| Should they be combined with other devices assessing complementary data (e.g., heart rate, O2, cardiopulmonary coupling (CPC), etc. | **1 2 3 4 (N/A)** |
| How confident are you in the use of movement-based devices combining movement recordings with other types of data (e.g., heart rate, O2). | **1 2 3 4 (N/A)** |
| How confident are you in the validity of “research-quality” devices in cognitively unimpaired older adults? (e.g., Actigraph, GENEActiv, etc) | **1 2 3 4 (N/A)** |
| How confident are you in the validity of “commercially-available” devices in cognitively unimpaired older adults? (e.g., Fitbit, Apple Watch, etc) | **1 2 3 4 (N/A)** |
| How confident are you in the validity of “research-quality” devices in MCI/dementia patients? (e.g., Actigraph, GENEActiv, etc) | **1 2 3 4 (N/A)** |
| How confident are you in the validity of “commercially-available” devices in MCI/dementia patients? (e.g., Fitbit, Apple Watch, etc) | **1 2 3 4 (N/A)** |
| In your opinion, which actigraphy devices are best suited to the examination of ageing/dementia? | Please comment *(open field)* |
| **PSG and EEG-based wearables** | |
| How confident are you in the validity of new EEG-based wearables to replace full PSGs? | **1 2 3 4 (N/A)** |
| How confident are you in devices such as WatchPat to replace full PSG? | **1 2 3 4 (N/A)** |
| Should they be combined with other devices assessing complementary aspects (e.g., heart rate, O2… | **1 2 3 4 (N/A)** |
| How confident are you in using oximeters for use at home to assess OSA in participants with MCI/dementia | **1 2 3 4 (N/A)** |
| How confident are you in the validity of such devices in cognitively unimpaired older adults? | **1 2 3 4 (N/A)** |
| How confident are you in the validity of such devices in participants with MCI/dementia? | **1 2 3 4 (N/A)** |

4. **Section 4: Moving the field forward: future directions**
   4.1.1. **What further evidence is required to establish or discard sleep problems as a risk factor for dementia? *Please check up to 5 priority areas.***

- More continuous monitoring of sleep (e.g., several days of recording in a row)
- More studies with longitudinal follow-up
- More biomarker/basic science studies
- More studies using self-report questionnaires
- More epidemiological studies
- More studies in clinical samples
- More PSG studies
- More mechanistic EEG studies, for example examining sleep microarchitecture
- More robust clinical trial data on the effect of sleep treatments on cognition, AD biomarkers
- More robust clinical trial data on the effect of sleep treatments on dementia onset/prevention
- Other, please specify *(open field)*
- Comments *(open field)*
  4.1.2. **Please identify the most understudied (yet potentially highly relevant) sleep and circadian features in the context of human studies of aging and AD.**

***Answer options: 1: not important, 2: minimally important, 3: moderately important, 4: strongly important, N/A: unable to answer (Please select this option if you are unsure or feel that an item falls out of your expertise).***

| Global self-reported measures (e.g., PSQI) | **1 2 3 4 (N/A)** |
| --- | --- |
| Excessive daytime sleepiness (e.g., ESS) | **1 2 3 4 (N/A)** |
| OSA | **1 2 3 4 (N/A)** |
| Parasomnias/RBD | **1 2 3 4 (N/A)** |
| Sleep-related movement disorders (e.g., PLMD) | **1 2 3 4 (N/A)** |
| Hypersomnolence disorders | **1 2 3 4 (N/A)** |
| Insomnia | **1 2 3 4 (N/A)** |
| Actigraphy features generally | **1 2 3 4 (N/A)** |
| Actigraphy-defined sleep variability | **1 2 3 4 (N/A)** |
| Harmonisation of actigraphy methodologies | **1 2 3 4 (N/A)** |
| Circadian rhythms using gold-standard circadian outputs | **1 2 3 4 (N/A)** |
| Naps | **1 2 3 4 (N/A)** |
| Sleep cycles | **1 2 3 4 (N/A)** |
| Sleep duration | **1 2 3 4 (N/A)** |
| Sleep fragmentation | **1 2 3 4 (N/A)** |
| N1 sleep | **1 2 3 4 (N/A)** |
| N2 sleep | **1 2 3 4 (N/A)** |
| N3 sleep / slow wave sleep | **1 2 3 4 (N/A)** |
| REM sleep | **1 2 3 4 (N/A)** |
| Spectral analyses/qEEG | **1 2 3 4 (N/A)** |
| EEG connectivity/coupling | **1 2 3 4 (N/A)** |
| Sleep spindles | **1 2 3 4 (N/A)** |
| Slow waves | **1 2 3 4 (N/A)** |
| K complexes | **1 2 3 4 (N/A)** |
| Epilepsy spikes characteristics | **1 2 3 4 (N/A)** |
| Other, please specify *(open field + rating)* | **1 2 3 4 (N/A)** |

4.1.3. **Please identify the most understudied populations in the context of aging and dementia.**

***Answer options: 1: not important, 2: minimally important, 3: moderately important, 4: strongly important, N/A: unable to answer (Please select this option if you are unsure or feel that an item falls out of your expertise).***

| Stratification according to biological sex | **1 2 3 4 (N/A)** |
| --- | --- |
| Individuals with genetic risk factors for AD (e.g., APOE ε4 carriers) | **1 2 3 4 (N/A)** |
| Extreme chronotypes | **1 2 3 4 (N/A)** |
| Short sleepers (<6h) | **1 2 3 4 (N/A)** |
| Long sleepers (>9h) | **1 2 3 4 (N/A)** |
| People with sleep disorders | **1 2 3 4 (N/A)** |
| Individuals with shift work history | **1 2 3 4 (N/A)** |
| Individuals with mental health disorders | **1 2 3 4 (N/A)** |
| Ethnical disparities | **1 2 3 4 (N/A)** |
| Non-AD dementias | **1 2 3 4 (N/A)** |
| People in low and middle income countries | **1 2 3 4 (N/A)** |
| Middle aged groups (40-65 years old) | **1 2 3 4 (N/A)** |
| Older groups (>65 years old) | **1 2 3 4 (N/A)** |
| Other, please specify *(open field + rating)* | **1 2 3 4 (N/A)** |

4.1.4. **Please identify the major future directions in the field of sleep and circadian rhythms research in the context of aging and AD.**

***Answer options: 1: not important, 2: minimally important, 3: moderately important, 4: strongly important, N/A: unable to answer (Please select this option if you are unsure or feel that an item falls out of your expertise).***

| Study sleep in under-studied and under-represented populations (e.g., ethnical disparities, sex and gender differences, other types of dementias, etc.) | **1 2 3 4 (N/A)** |
| --- | --- |
| Re-validate existing sleep questionnaires and potentially adapt their scoring to older populations with cognitive impairment | **1 2 3 4 (N/A)** |
| Develop a new sleep screening questionnaire specifically adapted to older people with cognitive impairment | **1 2 3 4 (N/A)** |
| Establish normative sleep data for older populations (for architecture, and microstructure/qEEG). | **1 2 3 4 (N/A)** |
| Identify critical/optimal time windows within which sleep disturbance should be identified to optimize brain health | **1 2 3 4 (N/A)** |
| Better understand the functions of sleep (e.g., glymphatics, cognition…) | **1 2 3 4 (N/A)** |
| Study the impact of lifestyle factors (e.g., physical activity) on sleep. | **1 2 3 4 (N/A)** |
| Gather more evidence on the treatment of sleep disorders/disturbances as a way to slow cognitive decline and/or impact the accumulation of AD pathology. | **1 2 3 4 (N/A)** |
| Determine if sleep should be screened for in dementia diagnosis | **1 2 3 4 (N/A)** |
| Determine if sleep should be screened in people at risk for AD (e.g., preclinical and prodromal AD) | **1 2 3 4 (N/A)** |
| Determine whether sleep interventions work in people at risk for AD (e.g., with cognitive impairment, risk factors) | **1 2 3 4 (N/A)** |
| Determine the role of circadian misalignment in AD pathology | **1 2 3 4 (N/A)** |
| Develop circadian treatments | **1 2 3 4 (N/A)** |
| Develop a global consortium for sleep and dementia data sharing | **1 2 3 4 (N/A)** |
| Other, please comment | (open field) |

4.2. **Do you routinely collect sleep data?**
- No
- Yes
  4.2.1. **Which type of data do you collect:**
- Self-report
- Actigraphy
- PSG
- Other, please specify
  4.2.2. **Would you be interested in future efforts to establish harmonized data consortia?**
- Yes
- No

*If yes, please provide contact details:*

- First and last name
- Email
- Affiliation
  4.2.3. **Do you consent to be contacted for the next iteration of this Delphi survey?**
- Yes
- No

*If yes, please provide contact details:*

- First and last name
- Email
- Affiliation
