## Appendix B for "International recommendations for sleep and circadian research in aging and Alzheimer’s disease: a Delphi consensus study"

CONSENSUS RECOMMENDATIONS ON RELEVANT SLEEP AND CIRCADIAN RHYTHMS MEASURES IN THE CONTEXT OF AGING AND ALZHEIMER’S DISEASE – **DELPHI SURVEY (ROUND 2).**

1. **Section 1: General information**
   1.1 Name
   1.2 Email address
   1.3 Primary affiliation(s)/Institution
   1.4 Country
   1.5 Postcode
   1.6 Professional setting *(Please tick all that apply)*

- Clinician
  - Neurologist
  - Geriatrician
  - Psychiatrist
  - Pulmonary physicians
  - Otolaryngologists
  - Anesthesiologists
  - General Practitioner (GP)/Primary Care Provider (PCP)
  - Neuropsychologist
  - Nurse practitioner (NP)
  - Physician assistant (PA)
  - Clinical psychologist
  - Occupational therapist
  - Sleep physician
  - Sleep technician
  - Other, *please specify: (open field)*

1.6.1	Are you still in clinical practice? Yes/No

1.6.2	Years in clinical practice, *please specify: (open field)*

- Researcher
  - Sleep medicine
  - Sleep neurophysiology
  - Alzheimer’s and dementia biomarkers,
  - …choose

Fluid biomarkers

Genetics

Imaging

- Epidemiology/Public health
- Health services
- Basic science
- Clinical or applied research
- Clinical trials
- Engineering or computer science
- Data science/AI
- Industry : Sleep technology company
- Industry : Pharmaceutical company
- Industry : Other
- Other research field, *please specify (open field)*

1.6.3	Are you still conducting research? Yes/No

1.6.4	Years of research experience, *please specify: (open field)*

*1.6.5	Are you an early career researcher (defined as <8 years since PhD)? Yes/No.*

1.7 Do you have any direct or indirect conflicts of interest relevant to the scope of this work (include consultancies, honoraria, company shares, or employment in industry)?

Yes/No

If Yes, please describe all relevant conflicts of interest hereafter *(open field)*

| Consultancies | *Open field* |
| --- | --- |
| Honoraria | *Open field* |
| Company shares | *Open field* |
| Employment in industry | *Open field* |
| Other, please specify | *Open field* |

1.8 Highest qualification
- Masters degree
- Medical degree (MD)
- PhD or equivalent
- Medical degree and PhD (e.g., MD, PhD)
- Medical degree and Masters degree (e.g., MD, MSc)
- Certified sleep technologist
- Other, *please specify (open field)*
1.9 Current primary position
    - PhD student/candidate
    - Postdoctoral Fellow
    - Faculty Academic appointment (e.g., Dr, Asst/Prof, Assoc.Prof, Prof) or full-time/tenured researcher
    - Clinical/medical hospital appointment (medical staff)
    - Director of institute
    - Company director
    - Employee of company/industry
    - Patient and public sleep advocacy group
    - Retired *(please specify your past primary position in the open field below)*
    - Other, *please specify (open field)*
1.10 Working environment *(check all that apply)*
     - Hospital or health service (outpatient facility)
     - Primary care
     - Private practice
     - Medical school
     - University
     - Research center or institute
     - Industry
     - Other, *please specify (open field)*
1.11 Are you from a low and middle income country? *Yes/No*
1.12 What is your sex
- Female
- Male
- Prefer not to answer
- Prefer to self describe (open field)
  1.13 What is your race/ethnicity?
       - American Indian or Alaskan Native
       - Asian
       - Black/African American
       - Hispanic/Latino
       - Native Hawaiian or Pacific Islander
       - White
       - Two or more races or ethnicities
       - Prefer not to answer
       - Prefer to self-describe

*Please self-describe: (open field)*

1.14 What is your primary language? *(open field)*
1.15 What is your secondary language? *(open field)*

2. **Section 2: Relevant sleep and circadian features**
   2.1. **Relevant features to characterize pathological ageing**
        2.1.1. **Please select the 10 measures which are the most sensitive to preclinical AD, among the following:**

NB: “Preclinical AD” here refers to cognitively unimpaired individuals who are engaged in the Alzheimer’s continuum (i.e., amyloid-positive at a minimum).

| *Variable* | *Measures sensitive to* ***preclinical AD*** |
| --- | --- |
| *Clinical and self-reports* |  |
| General sleep disturbance/poor sleep quality | ☐ |
| Insomnia | ☐ |
| Self-reported total sleep time (TST) | ☐ |
| Self-reported sleep efficiency (SE) | ☐ |
| *Polysomography* |  |
| Total sleep time (TST) | ☐ |
| Sleep efficiency (SE) | ☐ |
| Sleep fragmentation (e.g., number of awakenings, arousals, stages shifts) | ☐ |
| Wake after sleep onset (WASO) | ☐ |
| NREM sleep spectral power (including slow wave activity) | ☐ |
| Sleep spindles characteristics | ☐ |
| Slow waves characteristics | ☐ |
| EEG coupling/connectivity meaures | ☐ |
| *Actigraphy* |  |
| Total sleep time (TST) | ☐ |
| Sleep efficiency | ☐ |
| Wake after sleep onset (WASO) | ☐ |
| Daytime activity, including potential napping | ☐ |
| Other features derived from non-parametric or cosinor analysis | ☐ |

2.1.2. **Please select the 10 measures which are the most sensitive to MCI, among the following:**

| *Variable* | *Measures sensitive to MCI* |
| --- | --- |
| *Clinical and self-reports* |  |
| General sleep disturbance/poor sleep quality | ☐ |
| Sleep-related breathing disorders (e.g., OSA) symptoms | ☐ |
| Insomnia | ☐ |
| Self-reported sleep onset latency (SOL) | ☐ |
| Self-reported total sleep time (TST) | ☐ |
| Self-reported sleep efficiency (SE) | ☐ |
| Self-reported nocturnal awakenings (e.g., number or duration) | ☐ |
| Self-reported napping | ☐ |
| *Polysomnography/EEG* |  |
| Sleep onset latency (SOL) | ☐ |
| Total sleep time (TST) | ☐ |
| Sleep efficiency (SE) | ☐ |
| Sleep fragmentation (e.g., number of awakenings, arousals, stages shifts) | ☐ |
| Wake after sleep onset (WASO) | ☐ |
| Duration in each sleep stage/% | ☐ |
| Apnea-Hypopnea Index (AHI) | ☐ |
| Oxygen Desaturation Index (ODI) | ☐ |
| NREM sleep spectral power (including slow wave activity) | ☐ |
| REM sleep spectral power (including REM sleep EEG slowing) | ☐ |
| Sleep spindles characteristics | ☐ |
| Slow waves characteristics | ☐ |
| K complexes | ☐ |
| EEG coupling/connectivity meaures | ☐ |
| *Actigraphy* |  |
| Sleep onset latency (SOL) | ☐ |
| Total sleep time (TST) | ☐ |
| Sleep onset and offset | ☐ |
| Sleep efficiency | ☐ |
| Wake after sleep onset (WASO) | ☐ |
| Daytime activity, including potential napping | ☐ |
| Other features derived from non-parametric or cosinor analysis | ☐ |

2.1.3. **Please select the 10 measures which are the most sensitive to AD dementia, among the following:**

| *Variable* | *Measures sensitive to AD dementia* |
| --- | --- |
| *Clinical and self-reports* |  |
| General sleep disturbance/poor sleep quality | ☐ |
| Excessive daytime sleepiness | ☐ |
| Sleep-related breathing disorders (e.g., OSA) symptoms | ☐ |
| Insomnia | ☐ |
| Sleep timing (i.e., advanced or delayed sleep phase) | ☐ |
| Self-reported napping | ☐ |
| *Polysomnography/EEG* |  |
| Sleep onset latency (SOL) | ☐ |
| Total sleep time (TST) | ☐ |
| Sleep efficiency (SE) | ☐ |
| Sleep fragmentation (e.g., number of awakenings, arousals, stages shifts) | ☐ |
| Wake after sleep onset (WASO) | ☐ |
| Duration in each sleep stage/% | ☐ |
| Apnea Hypopnea Index (AHI) | ☐ |
| Oxygen Desaturation Index (ODI) | ☐ |
| NREM sleep spectral power (including slow wave activity) | ☐ |
| REM sleep spectral power (including REM sleep EEG slowing) | ☐ |
| Sleep spindles characteristics | ☐ |
| Slow waves characteristics | ☐ |
| K complexes | ☐ |
| EEG coupling/connectivity meaures | ☐ |
| *Actigraphy* |  |
| Sleep onset latency (SOL) | ☐ |
| Total sleep time (TST) | ☐ |
| Sleep onset and offset | ☐ |
| Sleep efficiency | ☐ |
| Wake after sleep onset (WASO) | ☐ |
| Daytime activity, including potential napping | ☐ |
| Other features derived from non-parametric or cosinor analysis | ☐ |

2.1.4. **How many days of actigraphy recordings are (i) the minimum standard and (ii) ideal to measure sleep and/or circadian rest-activity in a research setting?**

|  | **Sleep** | **Circadian rest-activity** |
| --- | --- | --- |
| Minimum standard | - <7 days - Between 7 and 13 days - ≥14 days - Unable to answer | - <7 days - Between 7 and 13 days - ≥14 days - Unable to answer |
| Ideal recording duration | - <7 days - Between 7 and 13 days - ≥14 days - Unable to answer | - <7 days - Between 7 and 13 days - ≥14 days - Unable to answer |

3. **Section 3: Recommendations on data acquisition and repor**t
   3.1. **Which method do you feel is best to assess the following features in each clinical group?**

| **Measure** | **Preclinical AD** | **MCI** | **AD dementia** |
| --- | --- | --- | --- |
| Time in Bed (TIB) | - Informant self-report - Participant self-report - Accelerometer / wearable devices - PSG/EEG | - Informant self-report - Participant self-report - Accelerometer / wearable devices - PSG/EEG | - Informant self-report - Participant self-report - Accelerometer / wearable devices - PSG/EEG |
| Sleep duration/total sleep time (TST) | - Informant self-report - Participant self-report - Accelerometer / wearable devices - PSG/EEG | - Informant self-report - Participant self-report - Accelerometer / wearable devices - PSG/EEG | - Informant self-report - Participant self-report - Accelerometer / wearable devices - PSG/EEG |
| Naps (timing, duration, frequency) | - Informant self-report - Participant self-report - Accelerometer / wearable devices - PSG/EEG | - Informant self-report - Participant self-report - Accelerometer / wearable devices - PSG/EEG | - Informant self-report - Participant self-report - Accelerometer / wearable devices - PSG/EEG |

3.2. **Do you think it is preferable to perform PSGs in the lab or at home?**

|  | **Preclinical AD** | **MCI** | **Dementia** |
| --- | --- | --- | --- |
| In-lab preferred | ◻︎ | ◻︎ | ◻︎ |
| At-home preferred | ◻︎ | ◻︎ | ◻︎ |
| Both are equally acceptable | ◻︎ | ◻︎ | ◻︎ |

3.3. **Consensus on the minimum standards for data collection and analysis in future studies/clinical trials.**
     3.3.1. **Recruitment and characterization of participants:** Please select the 5 most important demographic, clinical and biological features to assess:

| **Feature** | **5 most important features** |
| --- | --- |
| Gender | ☐ |
| Cognitive diagnosis | ☐ |
| Subjective cognitive/memory concerns | ☐ |
| Biomarker profile for clinical staging or ATN classification | ☐ |
| Race | ☐ |
| Ethnicity | ☐ |
| Education level | ☐ |
| Living arrangement/ Residential setting | ☐ |
| APOE ε4 | ☐ |
| Body mass index | ☐ |
| Lifestyle: physical activity levels | ☐ |
| Coffee, tea, energetic drinks consumption | ☐ |
| Daily light exposure | ☐ |
| Sleeping arrangement | ☐ |
| Menopause: current status | ☐ |

3.3.2. **Recruitment and characterization of participants:** Please select the 5 most important comorbidities (current and/or history) to assess:

| **Feature** | **5 most important features** |
| --- | --- |
| Smoking | ☐ |
| Alcohol intake | ☐ |
| Cannabis/THC | ☐ |
| Illicit substances | ☐ |
| State anxiety | ☐ |
| Trait anxiety | ☐ |
| Depressive symptoms | ☐ |
| Cardiovascular diseases | ☐ |
| Neurological diseases (e.g., stroke, epilepsy, Parkinsons) | ☐ |
| Psychiatric disorders and history of mental illness | ☐ |
| Metabolic syndromes/disorders | ☐ |
| Traumatic Brain Injury | ☐ |

3.3.3. **Recruitment and characterization of participants:** Please select the 5 most important sleep features to assess:

| **Feature** | **5 most important features** |
| --- | --- |
| Chronotype | ☐ |
| History of shift work | ☐ |
| Current shift work | ☐ |
| OSA diagnosis | ☐ |
| OSA treatment (e.g., CPAP use) | ☐ |
| Insomnia symptoms | ☐ |
| Insomnia treatment (e.g., CBT-I) | ☐ |
| Restless legs syndrome | ☐ |

3.3.4. **Recruitment and characterization of participants:** Please select the 5 most important medication type to assess:

| **Feature** | **5 most important features** |
| --- | --- |
| Benzodiazepines | ☐ |
| Other benzodiazepine receptor agonist hypnotics | ☐ |
| Trazadone | ☐ |
| Other approved hypnotics (e.g., DORAs, ramelteon…) | ☐ |
| Antidepressants. | ☐ |
| Antipsychotics | ☐ |
| Opioids | ☐ |
| Antihistamines | ☐ |
| Acetylcholinesterase (ACE) inhibitors | ☐ |
| Memantine | ☐ |
| Monoclonal antibody | ☐ |
| Non-narcotic analgesics | ☐ |
| Narcotic analgesics | ☐ |
| Anticonvulsants | ☐ |
| Stimulants (e.g., Ritalin) | ☐ |
| Hormone therapy | ☐ |
| Melatonin | ☐ |

3.3.5. **For cohort studies using polysomnography, do you feel a habituation night should be systematically performed?**
- No
- Yes, always (regardless of the clinical group)
- Only in cognitively unimpaired participants, but not MCI/dementia patients
- Unable to answer
  3.4. **Confidence in the use of newly available wearable and nearable devices.**

**Note:** In this section you will be assessing your confidence in recently developed wearable and "nearable" devices for sleep diagnostics and monitoring, encompassing both research-grade and commercially-accessible options.

- « Research quality » hereafter stands for medical grade and validated devices.
- « Consumer-based » defines devices that are commercially available but not approved by regulatory authorities as medical devices.
  3.4.1. For research purposes: How confident are you in accelerometer-based devices recording movement only to measure the following features?

| **Features** | **Research-quality accelerometer devices**  (e.g., Philips Actiwatch, ActigraphwGT3X-BT, GeneActiv, ActTrust) | **Consumer-based accelerometer devices**  (e.g., Oura ring, Evie Ring, Apple Watch, WHOOP, Fitbit, Garmin) |
| --- | --- | --- |
| Sleep onset latency (SOL) | - Confident - Not confident | - Confident - Not confident |
| Total sleep time (TST) | - Confident - Not confident | - Confident - Not confident |
| Sleep onset and offset | - Confident - Not confident | - Confident - Not confident |
| Sleep efficiency (SE) | - Confident - Not confident | - Confident - Not confident |
| Wake after sleep onset (WASO) | - Confident - Not confident | - Confident - Not confident |
| Daytime activity, including napping | - Confident - Not confident | - Confident - Not confident |
| Circadian features | - Confident - Not confident | - Confident - Not confident |

3.4.2. For research purposes: How confident are you in the validity of new EEG-based wearables to replace full PSGs?

| **Research-quality EEG devices**  (e.g., Dreem, Sleep profiler) | **Consumer-based EEG devices** (e.g., , Sleep profiler) |
| --- | --- |
| - Confident | - Confident |
| - Not confident | - Not confident |

3.4.3. For research purposes: How confident are you in the validity of wearable and nearable devices in MCI/AD dementia?

|  | **Research-quality devices**  (e.g., Dreem, Sleep Profiler,SleepImage, Accurable, Philips Actiwatch, wGT3X-BT actigraph) | **Consumer-based devices**  (e.g., , Fitbit, Apple Watch, Oura ring, Withings Sleep Tracking Mat, WHOOP, Muse S headband, Garmin Vivosmart 4). |
| --- | --- | --- |
| *Confident* | ☐ | ☐ |
| *Not Confident* | ☐ | ☐ |

4. **Section 4: Moving the field forward: future directions**
   4.1.1. **Please select the 5 features which are 1) the most important, which require more evidence, and 2) the most understudied to date yet potentially relevant in the context of AD.** *Please note that these categories are mutually exclusive, and a feature cannot be selected as “most important” and “most understudied” at the same time.*

| **Feature** | **Most important, and require further research** | **Most understudied to date, yet potentially relevant.** |
| --- | --- | --- |
| Global self-reported measures (e.g., PSQI) | ☐ | ☐ |
| Excessive daytime sleepiness (e.g., ESS) | ☐ | ☐ |
| OSA | ☐ | ☐ |
| Parasomnias/RBD | ☐ | ☐ |
| Sleep-related movement disorders (e.g., PLMD) | ☐ | ☐ |
| Hypersomnolence disorders | ☐ | ☐ |
| Insomnia | ☐ | ☐ |
| Actigraphy features generally | ☐ | ☐ |
| Actigraphy-defined sleep variability | ☐ | ☐ |
| Harmonisation of actigraphy methodologies | ☐ | ☐ |
| Circadian rhythms using gold-standard circadian outputs | ☐ | ☐ |
| Naps | ☐ | ☐ |
| Sleep cycles | ☐ | ☐ |
| Sleep duration | ☐ | ☐ |
| Sleep fragmentation | ☐ | ☐ |
| N1 sleep | ☐ | ☐ |
| N2 sleep | ☐ | ☐ |
| N3 sleep / slow wave sleep | ☐ | ☐ |
| REM sleep | ☐ | ☐ |
| Spectral analyses/qEEG | ☐ | ☐ |
| EEG connectivity/coupling | ☐ | ☐ |
| Sleep spindles | ☐ | ☐ |
| Slow waves | ☐ | ☐ |
| K complexes | ☐ | ☐ |
| Epilepsy spikes characteristics | ☐ | ☐ |

4.1.2. **Please select the 5 most understudied populations in the context of aging and dementia, which should be studied in priority in future research.**

| **Group** | **Rank** |
| --- | --- |
| Stratification according to biological sex | ☐ |
| Individuals with genetic risk factors for AD (e.g., APOE ε4 carriers) | ☐ |
| Extreme chronotypes | ☐ |
| Short sleepers (<6h) | ☐ |
| Long sleepers (>9h) | ☐ |
| People with sleep disorders | ☐ |
| Individuals with shift work history | ☐ |
| Individuals with mental health disorders | ☐ |
| Ethnical disparities | ☐ |
| Non-AD dementias | ☐ |
| People in low- and middle-income countries | ☐ |
| Middle aged groups (40-65 years old) | ☐ |
| Older groups (>65 years old) | ☐ |
